# Predicting Frequent Emergency Department Visitors using Adaptive Ensemble Learning Model

**DOI:** 10.1101/2024.10.31.24316535

**Authors:** Mehdi Neshat, Nikhil Jha, Michael Phipps, Chris A. Browne, Walter P. Abhayaratna

## Abstract

**Background:** Predicting accurately the frequent Emergency Department (ED) visitors is critical for hospitals because they often consume significant ED resources, including staff time, equipment, and medical supplies. Furthermore, frequent ED visitors may contribute to increased wait times for all patients. Therefore, by accurately predicting and identifying these individuals, hospitals can help reduce the burden on the ED and decrease wait times for all patients, improving the overall quality of care.

**Objective:** This study proposed an effective and adaptive ensemble learning prediction model to identify frequent visitors in the emergency department.

**Methods:** This was a retrospective population-based study of patients and utilised medical and administrative databases at Canberra Hospital, a tertiary public hospital in ACT, Australia, between January 1997 and December 2022. The study focuses on a wide age range of the population with 20 viral chronic diseases. The definition of frequent ED use is considered as having at least three visits within a year. This study developed an Adaptive ensemble learning–based prediction model and compared the performance with 16 popular machine learning models. In addition, three techniques are compared to handle the imbalanced data issue, and we also proposed a hybrid feature selection composed of Elastic-Net and local search to find the best combination of features. In order to hyper-parameter tuning, two techniques were compared: a population-based evolutionary algorithm and a local search.

**Results:** The study included 535,474 patient visits and 1.6 million episodes, with 25% overall frequent visitors. We compared the performance of the proposed prediction model with that of the other 16 popular classifiers. According to the prediction results, the proposed model considerably outperformed other models in terms of five metrics: accuracy, Recall, F1-score, Area under the ROC curve (AUC), and Log loss at 0.78 (95% CI 0.78-0.79), 0.68 (95% CI 0.68-0.68), 0.68 (95% CI 0.68-0.69), 0.69 (95% CI 0.69-0.70), and 7.4 (95% CI 7.2-7.5), respectively.

**Conclusions:** We proposed an adaptive ensemble learning model combining XGBoost Elastic-net with local search and Differential evolution to address the imbalanced nature of the frequent ED visitors’ data. Our approach aimed to enhance the prediction capability of the classifier substantially. To tackle the class imbalance, we employed both under-sampling and adjusted weights for the positive class. Through extensive testing and evaluation, we demonstrated that these strategies effectively improved the model’s performance. Further, we emphasised the importance of employing a robust feature selection method and a fast hyper-parameter optimiser. These elements were essential for enhancing the identification of frequent ED visitors. By incorporating these techniques, our study contributes to developing more accurate and reliable models for predicting frequent ED visitors, thereby assisting hospitals in resource allocation and patient care management.

## 1. Introduction

The Emergency Department (ED) is an essential component of hospitals as it fulfils multiple crucial roles [1]. It serves as a primary point of contact for immediate and life-saving care, ensuring round-the-clock availability to address emergent healthcare needs. The ED utilises a triage system to prioritize patient needs, facilitating efficient and timely care delivery. Definitions may vary, yet individuals who visit the ED at least thrice a year are categorised as “frequent users” [2]. These frequent ED visitors demonstrate a diverse array of characteristics, encompassing mental health issues, physical comorbidities, and disadvantaged socioeconomic status [3]. The requirements of these individuals are often too intricate to be adequately catered to within the confines of the ED environment [4]. Many of these recurrent ED visitors grapple with several chronic ailments, such as coronary artery disease or chronic obstructive pulmonary disease, conditions that could potentially be better managed in a primary care setting, thus averting acute deteriorations that necessitate ED visits [5]. The use of the ED to address complex needs points towards a less-than-optimal scenario where these needs have not been satisfactorily addressed within the realm of primary care. This form of ED use is linked with unfavourable outcomes for patients, including heightened rates of hospital admissions and mortality [6]. Additionally, the costs associated with ED services tend to be elevated in comparison to those in primary care, thereby imposing a socioeconomic strain on the healthcare system [7]. As an illustration, a particular study conducted in the province of Quebec, Canada, [2] revealed that individuals classified as frequent ED users with chronic diseases constituted approximately 9.2% of the total ED user population. Surprisingly, these frequent users were regarded as a significantly higher proportion, around 28.8%, of all ED visits. Another research [8] endeavour focused on defining frequent users as those with four or more ED visits annually. The findings indicated that this subset of patients represented approximately 4.5% to 8% of the overall ED patient population. Strikingly, they contributed to a substantial 21% to 28% of all ED visits. These statistics highlight frequent users’ significant impact and disproportionate utilisation of ED resources, emphasising the need for targeted interventions and alternative care strategies to address their specific needs and reduce the strain on the ED system.

Accurately identifying high-rate visitors among normal visitors is a crucial step in mitigating unnecessary ED visits. Extensive research has been conducted to tackle this challenge using statistical models [9]. Logistic regression (LR) has emerged as a standard and widely employed statistical model for this purpose [10]. Although LR is known for its effectiveness in prediction tasks, it encounters limitations when applied to predict frequent ED visitors. Its linear nature assumes a linear relationship between predictor variables and the target, which may not hold true for this case study. The relationship between predictors, such as demographic factors and medical history, and the target of being a frequent visitor often exhibit complexity and nonlinearity [11]. LR may struggle to accurately capture and model these intricate nonlinear associations, potentially compromising its predictive performance in this context.

Traditional statistical models face limitations in identifying highly visited ED patients due to their reliance on predetermined rules centred around specific clinical predictors. On the other hand, machine learning (ML) prediction models present a more adaptable approach through the utilisation of nonparametric algorithms that can encompass a broad spectrum of intricate predictors while upholding robust predictive capacities. The ML-based techniques application brings about advantages such as learning error reduction, runtime and cost savings, as well as an enhancement in the quality of care services provided [12]. Prior studies have focused on applying ML models in predicting frequent visitors, with numerous research works disseminating their discoveries in this domain [13]. This exploration has showcased the potential of ML in revolutionising the identification and management of frequent ED patients, marking a significant leap in the realm of healthcare analytics.

When predicting frequent visitors in the Emergency Department (ED), neural networks have gained significant attention among the various machine learning (ML) models utilised. This popularity can be attributed to their remarkable ability to learn and model intricate patterns and relationships within both small and large-scale datasets. Neural networks (NNs) encompass a range of architectures, including feed-forward backpropagation (FFNN) [2], multilayer perceptron (MLP) [14], deep neural networks (DNN) [15] and hybrid deep learning models [16]. These architectures enable neural networks to effectively capture the complexity of the data, making them well-suited for tasks involving frequent ED visitor prediction. Despite their remarkable capabilities, NNs are subject to various constraints. A significant drawback is the lack of interpretability. Moreover, NNs demand substantial computational power and resources, resulting in elevated computational and resource demands. Overfitting presents another issue: NNs may excessively specialise in the training data, leading to subpar generalisation. Lastly, identifying optimal hyper-parameters for NNs can be intricate, necessitating meticulous adjustments.

Recently, a notable surge of interest has been in utilising tree-based and ensemble learning models to predict frequent ED visitors. The Random Forest (RF) [2, 17, 18] model is one popular choice in related studies and is known for its robust classification capabilities [19]. Another extensively employed model is XGB [20], which excels in handling small datasets with complex patterns. Among the commonly utilised models, XGB often exhibits superior predictive performance [21]. However, it is not without its limitations, such as the need for hyperparameter tuning. To strike a favourable balance, some studies have adopted ensemble models like the voting classifier [14], which combines multiple models with specific weighting ratios. This approach allows for a more comprehensive and nuanced prediction by leveraging the strengths of different models within the ensemble.

In this study, we build upon previous research by developing a series of binary classifiers using a dataset consisting of 535,474 patient visits spanning 25 years at the ED of Canberra Hospital in ACT, Australia. For each visit, we extracted 18 variables, including patient demographics, primary diagnosis, up to 20 secondary diagnoses coded in ICD-10, admission and release dates, triage category, and patient disposition. To predict frequent ED visitors, we establish a comprehensive comparative framework employing 16 machine learning models. These models encompass linear and logistic regression, various types of neural networks, popular ensemble learning models, and deep learning models, enabling us to capture the nonlinear relationships among the variables. Furthermore, we introduce an adaptive feature selection approach by combining Elastic-Net and a local search technique to optimise the performance of our feature set. To fine-tune the best-performing model, we implement an effective hyper-parameter optimisation method utilising Differential Evolution (DE). Through this research, we aim to provide insights into predicting frequent ED visitors and enhance the overall understanding of patient visit patterns in the healthcare system.

## 2. Study design and data sources

In this population-based retrospective cohort study, data was collected from the Emergency Department of Canberra Hospital, which serves as the largest and oldest public hospital in Canberra, Australia’s ACT province. The collected period spans 25 years, from January 1997 to December 2022. The dataset encompasses various components, including the patient demographic register, which provides information such as sex, date of birth, and place of residence. Additionally, features extracted from the hospital register include the primary diagnosis based on the International Classification of Diseases, tenth revision (ICD-9), as well as up to 20 secondary diagnoses coded using ICD-10. Other relevant details encompass admission and release dates, triage category, and patient disposition. For further insights into the specific features utilised in this study, Supplementary Table 1 can be referred to.

**Table 1:** Descriptive statistics for the different populations of whole visitors and frequent visitors (3+ visits per Year) from 1997 to 2022. Percentages in brackets are relative to the column base.

| Variable |  | Total (%) | Non-frequent users 3 (%) | Frequent users 3 (%) |
| --- | --- | --- | --- | --- |
| Total population |  | 535474 (100) | 475875 (100) | 59599 (100) |
| Female |  | 255869 (47.78) | 226885 (47.68) | 28984 (48.63) |
| Age | <=18 | 152901 (**28.55**) | 138727 (29.15) | 14174 (**23.78**) |
|  | 18-25 | 60875 (11.37) | 54691 (11.49) | 6184 (10.38) |
|  | 26-35 | 78560 (14.67) | 70272 (14.77) | 8288 (13.91) |
|  | 36-45 | 61351 (11.46) | 54832 (11.52) | 6519 (10.94) |
|  | 46-55 | 54136 (10.11) | 48602 (10.21) | 5534 (9.29) |
|  | 56-65 | 45966 (8.58) | 41083 (8.63) | 4883 (8.19) |
|  | 66-75 | 36749 (6.86) | 31858 (6.69) | 4891 (8.21) |
|  | >75 | 44861 (8.38) | 35735 (7.51) | 9126 (15.31) |
| Triage | 1 (Red) | 7243 (1.35) | 6545 (1.38) | 698 (1.17) |
|  | 2 (Orange) | 57188 (10.68) | 49256 (10.35) | 7932 (13.31) |
|  | 3 (Yellow) | 170228 (31.79) | 148645 (31.24) | 21583 (36.21) |
|  | 4 (Green) | 216441 (**40.41**) | 194565 (40.89) | 21876 (**36.70**) |
|  | 5 (Blue) | 84281 (15.74) | 76774 (16.13) | 7507 (12.60) |
| State | ACT | 391095 (73.04) | 338412 (71.11) | 52683 (88.40) |
|  | NSW | 121374 (22.67) | 114901 (24.15) | 6473 (10.86) |
| Suburb | Kambah (8.6 Km) | 20765 (**5.31**) | 17310 (5.12) | 3455 (**6.56**) |
|  | Wanniassa (9.2 Km) | 10496 (2.68) | 8684 (2.57) | 1812 (3.44) |
|  | Narrabundah (5.5 Km) | 9862 (2.52) | 8080 (2.39) | 1782 (3.38) |
|  | Monash (9.5 Km) | 7803 (1.96) | 6296 (1.86) | 1507 (2.86) |
|  | Gordon (15 Km) | 10250 (1.91) | 8494 (2.51) | 1756 (3.33) |
| Disposition | Admit | 184381 (34.43) | 159271 (33.47) | 25110 (42.13) |
|  | Home | 307159 (**57.36**) | 277290 (58.27) | 29869 (**50.12**) |
|  | Did not wait (DNW) | 30209 (5.64) | 27053 (5.68) | 3156 (5.30) |
|  | Left at own risk (LOR) before treatment completed | 1525 (0.28) | 1270 (0.27) | 255 (0.43) |
|  | Discharged to DHR | 40 (0.007) | 27 (0.01) | 13 (0.02) |
|  | Referred to other TCH service | 8079 (1.51) | 7422 (1.56) | 657 (1.10) |
|  | Transferred to other hospital | 2333 (0.44) | 2039 (0.43) | 294 (0.49) |
|  | Went to GP | 435 (0.44) | 423 (0.09) | 12 (0.02) |
|  | Died in ED | 972 (0.18) | 790 (0.17) | 182 (0.31) |

Two exclusion criteria were applied in this study, as depicted in Figure 1. The first criterion involved excluding patients with missing information, which accounted for a negligible rate of 0.004%. The second criterion entailed excluding patients who had more than three Emergency Department (ED) visits per year, which constituted a substantial portion of 89% of the total number of patients. By implementing these exclusion criteria, the study aimed to ensure data completeness and focus on individuals with a higher frequency of ED visits for a more comprehensive analysis.

**Figure 1:**
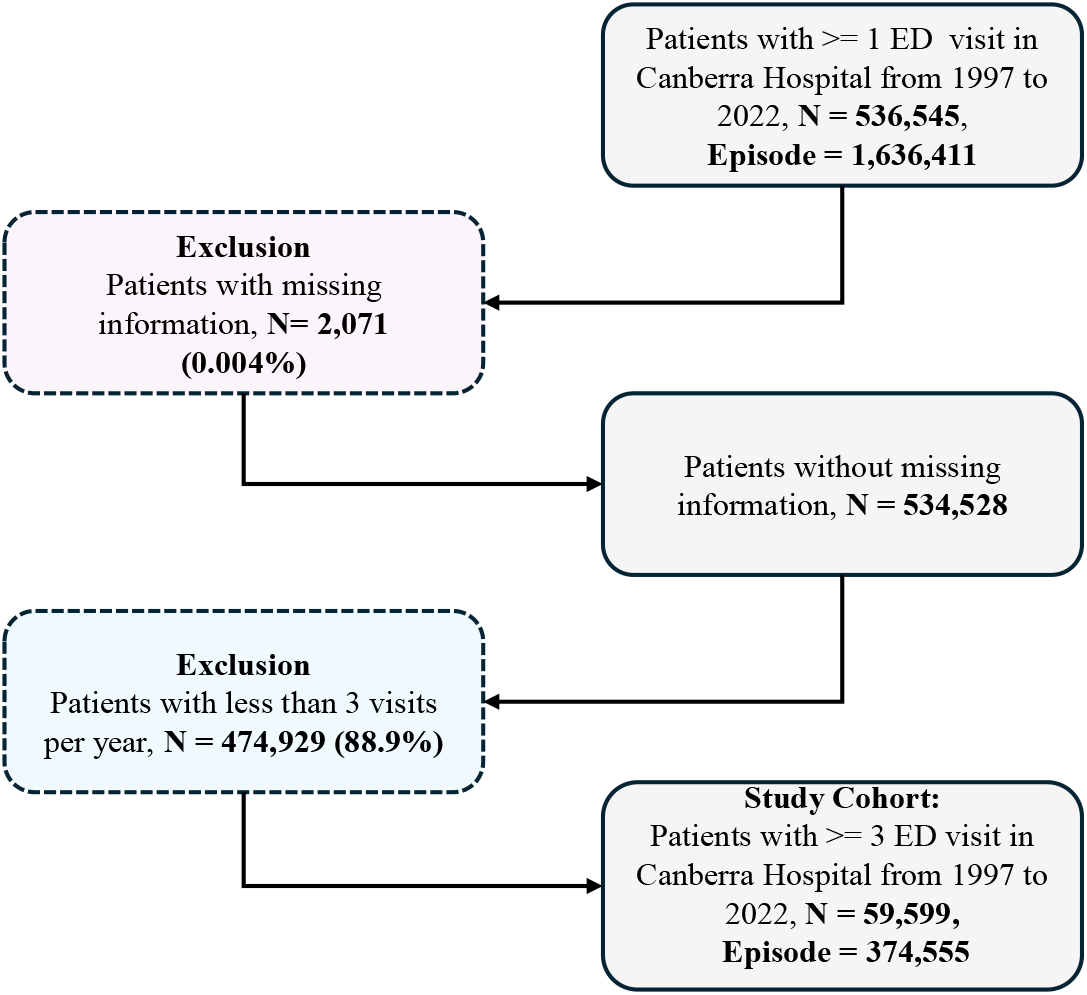
Flowchart of the study cohorts. The number of patients with ≥ 3 visits is 59,599, and the episode number is 374,555.

As reported, the number of ED visitors at Canberra Hospital has increased by an average of 4% over the years (See Figure 2). However, a notable dip was observed in 2020, reflecting a substantial decrease in ED visitors due to the implementation of the COVID-19 lockdown situation. Based on predictive analysis, it is estimated that the monthly average of ED episodes will reach 11,000 in 2032 and subsequently rise to 13,000 in 2042. These projections indicate a steady upward trend in ED visits over the next two decades, with the exception of the temporary decline observed in 2020.

**Figure 2:**
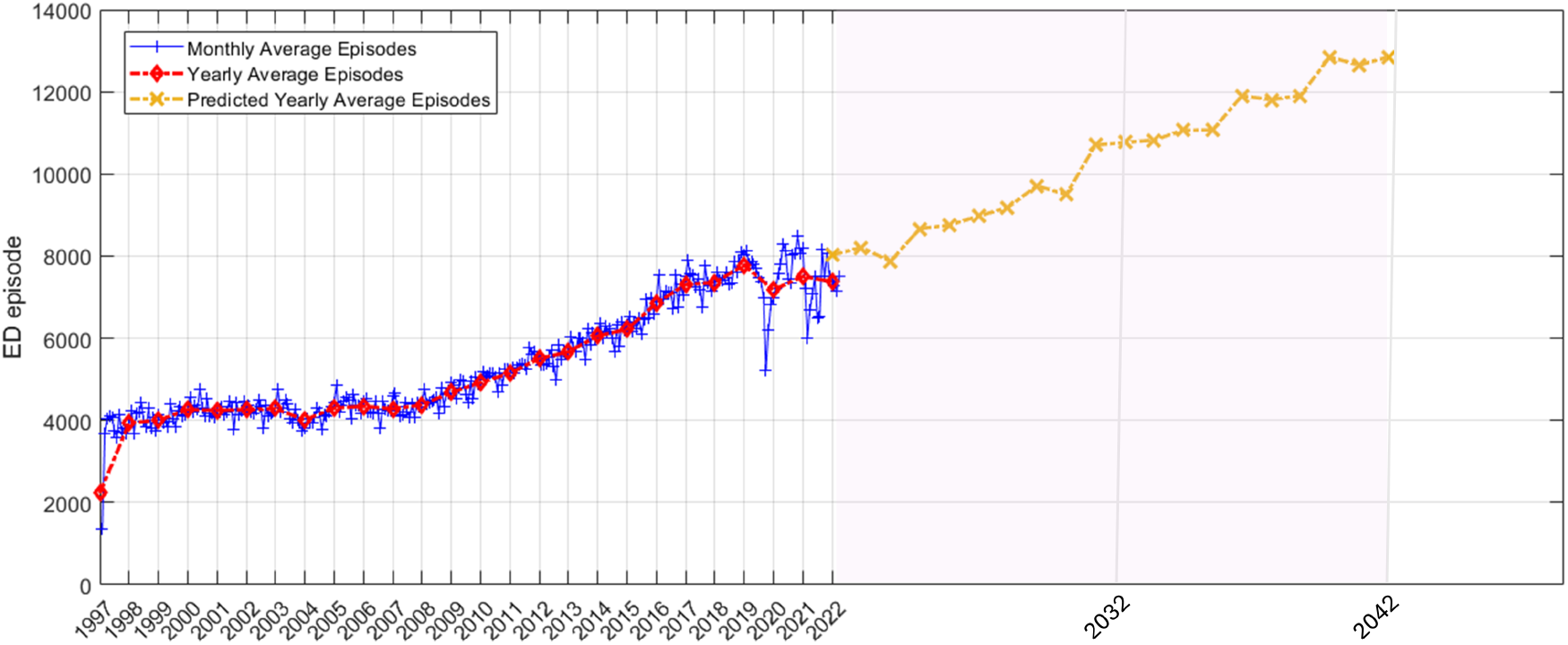
The monthly and annual average of ED visitors episodes in the Canberra Hospital from 1997 to 2022, as well as the predicted increase in the visitors for the next 20 years.

### 2.1 Characteristics of participants

Descriptive statistics for various populations, including whole ED visitors and frequent visitors (3+ visits per year), as well as information on triage category, patient location (state and suburb), and patient disposition, are presented in Table 1. The table highlights a significant finding: the largest proportion of non-visitors and frequent visitors is among individuals under 18 years of age, accounting for 29.15% and 23.78% of their respective populations. Furthermore, among frequent visitors, approximately half of the patients were discharged to their homes, indicating a common disposition. Additionally, it is notable that the triage category labelled as “green” exhibits the highest rate at 36.70% among the different triage categories, indicating a relatively lower level of urgency or severity in those cases.

A comprehensive statistical analysis was conducted on the monthly (Table 2) and weekday (Table 3) data of whole, non-, and frequent visitors at Canberra Hospital. The findings indicate that October and November emerge as the busiest months, accounting for 10.81% and 9.88% of the total high-frequency visitors, respectively. These peaks could be attributed to various factors, including the coinciding spring season in Australia, which brings about an allergy season, increased prevalence of respiratory infections, and a higher risk of seasonal asthma [22] exacerbations. Additional contributing factors may also play a role in the observed patterns. The distribution of non- and frequent ED visitors across the seven weekdays is presented in Table 3. The analysis reveals that Monday experiences the highest overload in the Canberra Hospital’s ED, accounting for 15.76% of the total visits. This finding suggests that Mondays exhibit a greater demand for emergency services compared to other weekdays.

**Table 2:** Monthly statistical analysis of whole ED visitors and frequent visitors (3+ visits per Year) from 1997 to 2022.

| Variable | Total (%) | Non-frequent users 3 (%) | Frequent users 3 (%) |
| --- | --- | --- | --- |
| Population | 535474 (100) | 475875 (100) | 59599 (100) |
| January | 41120 (7.68) | 37643 (7.91) | 3477 (5.83) |
| February | 39126 (7.31) | 35539 (7.47) | 3587 (6.01) |
| March | 44725 (8.35) | 40557 (8.52) | 4168 (6.99) |
| April | 42423 (7.92) | 38236 (8.03) | 4187 (7.03) |
| May | 45365 (8.47) | 40074 (8.42) | 5291 (8.88) |
| June | 43441 (8.11) | 38510 (8.09) | 4931 (8.27) |
| July | 46012 (8.59) | 40736 (8.56) | 5276 (8.85) |
| August | 47744 (8.92) | 42058 (8.83) | 5686 (9.54) |
| September | 46518 (8.69) | 40855 (8.58) | 5663 (9.50) |
| October | 48593 (**9.07**) | 42149 (8.85) | 6444 (**10.81**) |
| November | 45493 (8.50) | 39603 (8.32) | 5890 (9.88) |
| December | 44914 (8.39) | 39915 (8.39) | 4999 (8.39) |

**Table 3:** Weekday statistical analysis of whole ED visitors and frequent visitors (3+ visits per Year) from 1997 to 2022.

| Variable | Total (%) | Non-frequent users 3 (%) | Frequent users 3 (%) |
| --- | --- | --- | --- |
| Population | 535474 (100) | 475875 (100) | 59599 (100) |
| Saturday | 79049 (14.76) | 71265 (14.98) | 7784 (13.06) |
| Sunday | 80105 (**14.84**) | 71925 (15.11) | 8180 (13.73) |
| Monday | 79484 (13.99) | 70094 (14.73) | 9390 (**15.76**) |
| Tuesday | 74903 (13.99) | 66093 (13.89) | 8810 (14.78) |
| Wednesday | 73576 (13.74) | 64990 (13.66) | 8586 (14.41) |
| Thursday | 73673 (13.76) | 65113 (13.68) | 8560 (14.36) |
| Friday | 74684 (13.95) | 66395 (13.95) | 8289 (13.91) |

In order to gain insights into the primary reasons for ED visits, we conducted an analysis of viral diagnoses among both non- and highly frequent visitors. Table 4 presents a compilation of the top 25 viral diagnoses, sorted in descending order. Notably, Chest pain (R07.4), Abdominal Pain (T79.4), and Pain in the abdomen (including colic) (R10.4) emerged as the most prevalent diagnoses, accounting for rates of 3.9%, 3.44%, and 3.44%, respectively. These findings highlight the significance of these conditions and underscore the need for appropriate management and attention to these specific health concerns.

**Table 4:** Statistical analysis of most common causes to visit ED based on whole visitors and frequent visitors (3+ visits per Year) from 1997 to 2022.

| Variable | Total (%) | Non-frequent users 3 (%) | Frequent users 3 (%) |
| --- | --- | --- | --- |
| Population | 535474 (100) | 475875 (100) | 59599 (100) |
| Did not wait for treatment (Z53.1) | 22480 (**4.2**) | 20832 (4.38) | 1648 (2.77) |
| Chest pain, unspecified (R07.4) | 19011 (**3.11**) | 16698 (3.51) | 2313 (**3.88**) |
| Abdominal pain (T79.4) | 3649 (0.68) | 1601 (0.34) | 2048 (**3.44**) |
| Suicidal ideation (R45.81) | 3027 (0.56) | 2336 (0.49) | 691 (1.16) |
| Pain in abdomen, other (includes colic) (R10.4) | 16651 (**3.11**) | 14603 (3.07) | 2048 (**3.44**) |
| Viral infection, unspecified (B34.9) | 9465 (1.77) | 8485 (1.78) | 980 (1.64) |
| For review (Z09.9) | 9109 (1.70) | 7933 (1.67) | 1176 (1.97) |
| Injury, unspecified or suspected of head (S09.9) | 6125 (1.14) | 5630 (1.18) | 495 (0.83) |
| Open wounds, lacerations - finger - uncomplicated (S61.0) | 5345 (1.0) | 4992 (1.05) | 353 (0.59) |
| Urinary tract infection (N39.0) | 5068 (0.95) | 4768 (1.00) | 300 (0.50) |
| Syncope or collapse (except heat syncope) (R55) | 4882 (0.91) | 4412 (0.93) | 470 (0.79) |
| Sprain and strain of ankle, part unspecified (S93.40) | 4803 (0.90) | 4446 (0.93) | 357 (0.60) |
| Nausea and vomiting (except in pregnancy) (R11) | 4679 (0.87) | 4021 (0.84) | 658 (1.10) |
| Asthma, Acute (J45.9) | 4595 (0.86) | 3885 (0.82) | 710 (1.19) |
| Low back pain (M54.5) | 4241 (0.8) | 3726 (0.78) | 515 (0.86) |
| Headache (R51) | 3983 (0.74) | 3471 (0.73) | 512 (0.86) |
| Fever (R50.9) | 3947 (0.74) | 3430 (0.72) | 517 (0.87) |
| Pneumonia, unspecified (J18.9) | 3695 (0.69) | 2967 (0.62) | 728 (1.22) |
| Multiple trauma (T79.4) | 3649 (0.68) | 3586 (0.75) | 63 (0.10) |
| Upper respiratory tract infection (URTI), acute (J06.9) | 3548 (0.66) | 3111 (0.65) | 437 (0.73) |
| Gastroenteritis and colitis of unspecified origin (A09.9) | 3546 (0.66) | 3054 (0.64) | 492 (0.83) |
| Tonsillitis (J03.9) | 3354 (0.63) | 3005 (0.63) | 349 (0.59) |
| Stroke, not specified as haemorrhage or infarction (I64) | 1843 (0.34) | 1581 (0.33) | 262 (0.44) |
| Migraine (G43.9) | 1806 (0.34) | 1561 (0.33) | 245 (0.41) |
| Sepsis (except with notifiable agent) (A41.9) | 1904 (0.36) | 1419 (0.30) | 485 (0.81) |

A comprehensive analysis of all 1.6 million episodes in the ED at Canberra Hospital from 1997 to 2022, including frequent visitors with more than 3 and 5 visits per year, was conducted to identify the top 20 reasons for attending. Figure 3 showcases these findings, and remarkably, the most prevalent reason for seeking ED services was visitors who did not wait for medical attention, encompassing roughly 6% of the total episodes across all three groups. Subsequently, the second and third most common reasons were identified as Abdominal Pain and Chest Pain, respectively.

**Figure 3:**
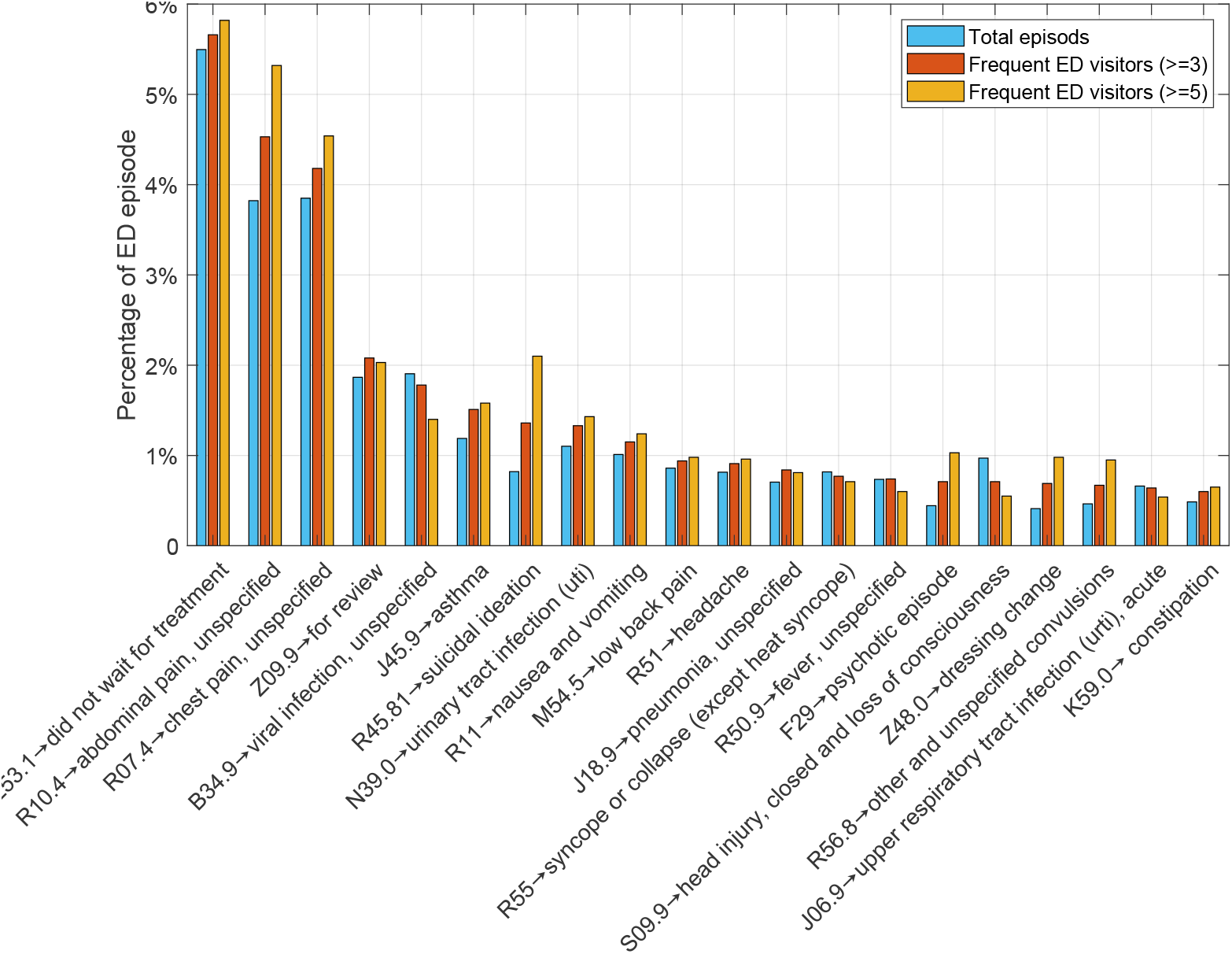
ED visits comparison by all episodes, frequent visitors with more than 3 and 5 visits per year sorted within 20 viral reason categories.

To gain a deeper understanding of the relationship between the age of frequent visitors and the 20 most common viral reasons for ED visits, we divided the patients into five age groups, as depicted in Figure 4(a). The analysis revealed distinct age patterns associated with viral reasons. Notably, patients aged ≤ 14 represented the highest proportion of ED visitors for Viral infection (B34.9), Asthma (J45.9), Head Injury (S09.9), Fever (R50.9), and Upper respiratory tract infection (J06.9), accounting for 60%, 59%, 49%, 58%, and 70% of the total episodes, respectively. Conversely, senior visitors (age ≥ 64 years) exhibited a different pattern, with Pneumonia (J18.9), Syncope (R55), Urinary tract infection (N39.0), and Constipation (K59.0) being the prominent viral reasons. These conditions were observed at rates of 50%, 50%, 39%, and 35%, respectively, among the other population. Having insights into the age-specific trends in viral reasons for ED visits can be helpful for healthcare professionals to tailor their approaches to meet the unique needs of different age groups. The seasonality analysis of the most common disorders among highly-frequent visitors is presented in Figure 4(b). The findings indicate that, apart from viral infection, pneumonia, and respiratory tract infection, which exhibit a notable pattern of being more prevalent during winter, no specific seasonal trend was observed for the other disorders. These three conditions demonstrate a higher frequency during the winter months, potentially influenced by factors such as increased viral circulation, colder temperatures, and reduced immunity.

**Figure 4:**
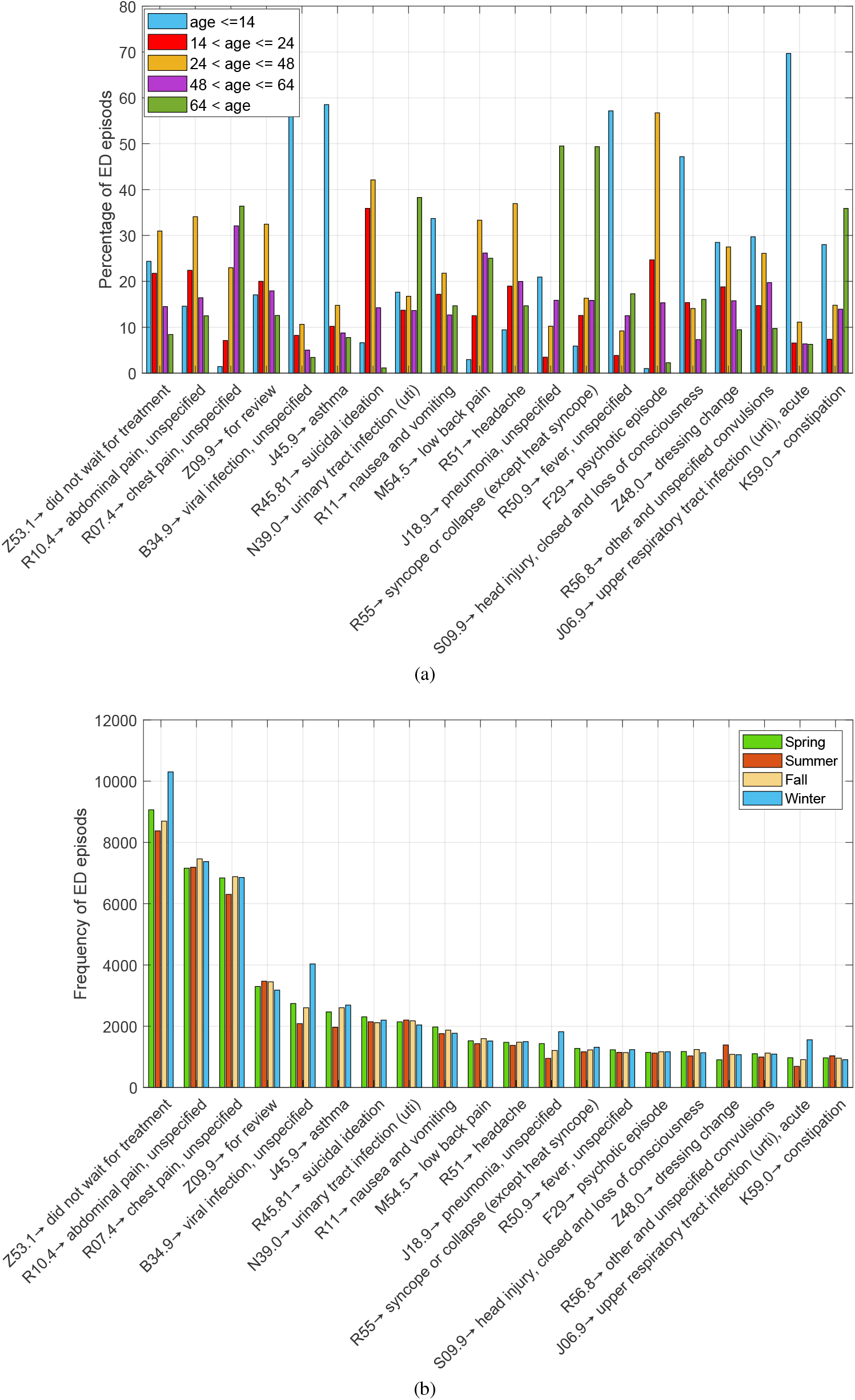
Age groups (a) and Seasonal ED visits(b) for frequent visitors with more than 3 visits per year sorted within 20 viral reason categories.

In Figure 5, a noteworthy pattern is evident concerning the gender distribution of frequent visitors in the 20 most prevalent reasons for ED visits. Specifically, there are significant gender differences in the contribution percentages for certain conditions. Among the identified conditions, it is observed that females account for a higher proportion in several categories. For instance, in Urinary tract infections, females contribute to ED visits at a rate of 137% compared to males. Similarly, for Abdominal pain, Headache, Suicidal ideation, Nausea and vomiting, and Low back pain, the female contribution percentages are 108.71%, 74.68%, 65.11%, 56.92%, and 24.77%, respectively. On the other hand, there is a noticeable trend of increased representation among males for specific disorders when compared to females. Conditions such as dressing changes, Head injuries, Psychotic episodes, Fever, and Pneumonia exhibit higher contribution rates among males, with percentages of 51.77%, 40.02%, 41.63%, 25.31%, and 34.66%, respectively.

**Figure 5:**
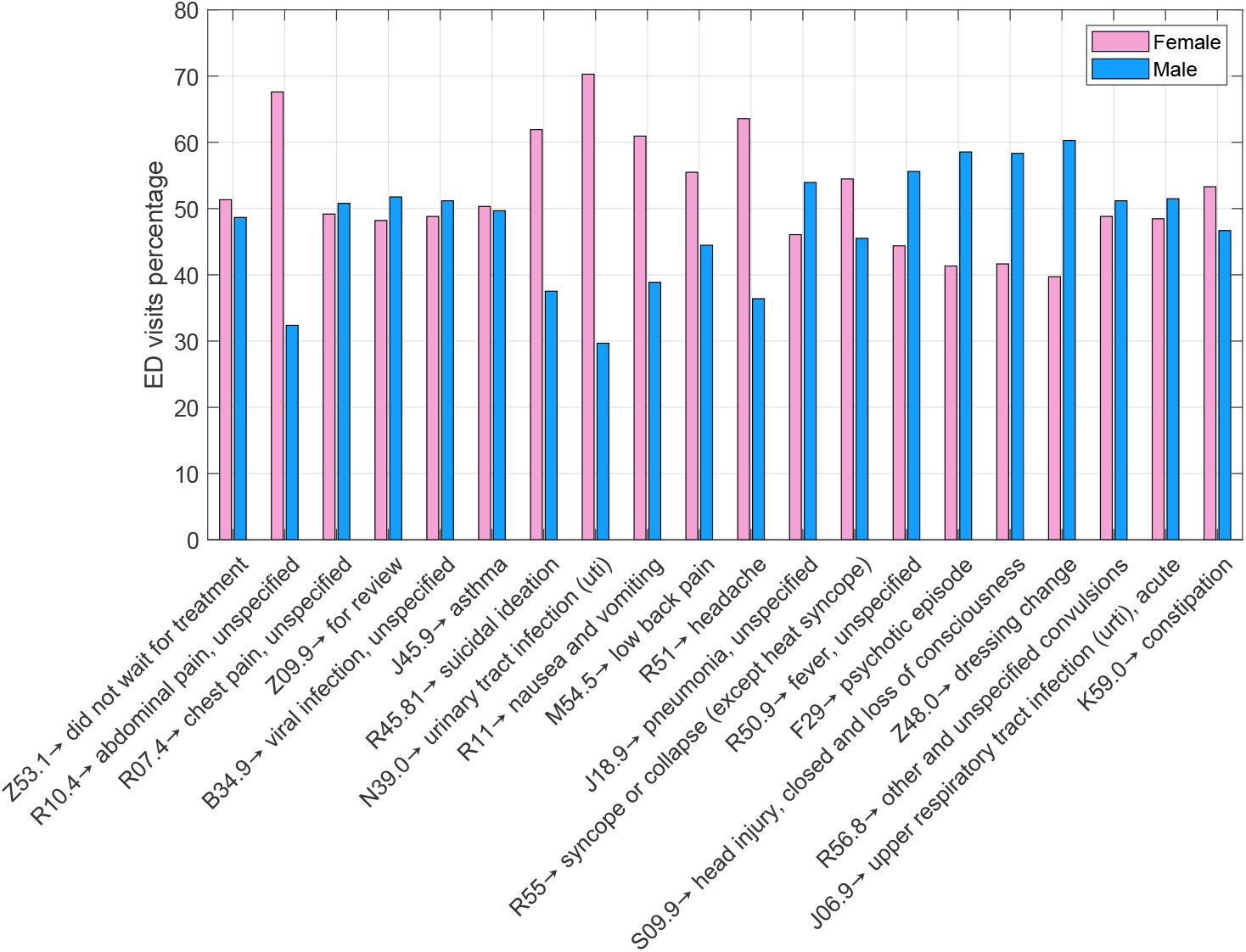
Gender groups for frequent visitors with more than 3 visits per year sorted within 20 viral reason categories.

## 3. Methods

### 3.1 Ensemble Machine learning models

In opposition to conventional data-centric modelling strategies that entail fitting a single predictive model, ensemble-based models amalgamate the forecasts of numerous base estimators to amplify resilience [23]. The ensemble learning techniques at the core encompass bagging [24], boosting [25], and stacking [26]. Bagging entails the random selection, with replacement, of multiple subsets of training data to train an identical model. The training of these models operates independently of one another and can be executed concurrently. The ultimate forecast is the mean of predictions generated by all models. Bagging enhances generalisation and mitigates overfitting. In contradistinction to bagging, the boosting methodology involves the sequential training of models, where each fresh model strives to refine predictions by concentrating on the feeble forecasts of preceding models within the ensemble. Boosting stands out as one of the most potent learning methodologies unveiled in the realm of machine learning [27]. Stacking emerges as a more intricate ensemble technique. Instead of directly averaging or sequentially amalgamating base model predictions, akin to bagging and boosting, stacking harnesses these individual base models’ predictions as input for a superior-level model. This meta-model is schooled on these varied base model predictions to craft precise final predictions. The essence of stacking lies in its ability to leverage the diversity among the base models to enhance the overall predictive accuracy.

### 3.2 Gradient Boosting Machine (GBM)

This innovative method follows a forward-thinking, gradual procedure where new cutting-edge predictive models are integrated into the group one by one. The process typically relies on regression trees (RTs) as the basic weak learners. These RTs are generated sequentially, aiming to capture any prediction mistakes left by the preceding RTs. The Gradient Boosting Machine [28] (GBM) begins by creating a differentiable loss function, often using the squared error as the loss function, which can be symbolized as a mathematical expression. The GBM then iterates through the ensemble, emphasising correcting the errors made by the previous models by adjusting the weights of each model based on the loss function’s gradient. This iterative process continues until a specified stopping criterion is met, producing a powerful ensemble model that combines the strengths of individual weak learners to make accurate predictions.

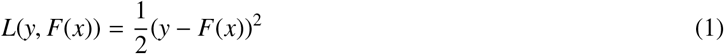

Within the domain of mathematical symbolism, the symbols *y* and *F* carry a unique and profound meaning, representing the actual output results and the forecasts produced by the less powerful learner. One search through the commencement phase involves reflecting on a fixed forecast value attributed to each individual data point, a particular computation that is executed in a subsequent manner:

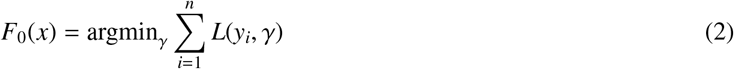

Suppose the decision is made to utilise Equation 1 as the designated loss function; the effects of Equation 2 will materialise in the form of a direct calculation representing the mean of the actual results associated with the training dataset. Following this, the subsequent steps entail performing calculations to determine pseudo-residuals for every single data point, a process that is executed by following a specific approach and methodology. This systematic approach involves analysing each data point to accurately compute the residual values, which is crucial in further refining the model’s performance and optimising the training process.

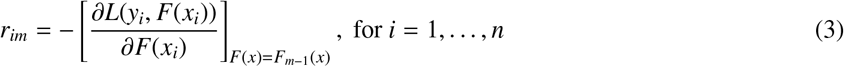

Delving deeper into the realm of advanced algorithms, the powerful and sophisticated GBM embarks on the challenging task of training a base learner, which can take the form of a tree or any other type of model, and then adjusts it in accordance with the pseudo-residuals meticulously crafted during the process. This particular base learner is symbolically represented as *F*_*m*_(*x*), with the training dataset 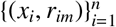 laying the essential groundwork for this intricate journey towards optimisation. As the journey progresses to the next phase, the GBM meticulously computes the multiplier ρ_*m*_ by carefully addressing the optimisation problem that is elegantly presented in a structured manner. The interplay between the base learners and the pseudo-residuals within the GBM framework showcases a harmonious blend of mathematical precision and algorithmic finesse, resulting in a dynamic and adaptive learning process that continually refines the model’s predictive capabilities. Each step in the GBM’s iterative process unfolds like a careful control, where each base learner contributes unique insights while guided by the significant objective of minimising the loss function. Through this intricate interplay of components and calculations, the GBM methodically enhances the model’s predictive power, unlocking new possibilities and insights in the realm of machine learning that is presented in the following manner:

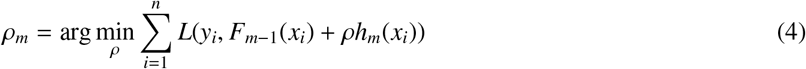

therefore, the model is updated as below:

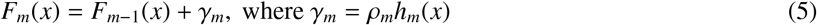

### 3.3. Extreme Gradient Boosting (XGBoost)

This innovative technique significantly improves the Gradient Boosting Machine (GBM) by incorporating novel regularization methods that effectively reduce the chances of overfitting and complex modeling [29]. The main objective of this advanced approach, as demonstrated in Equation 6, is to balance the training loss and model complexity, resulting in the development of streamlined prediction models with exceptional predictive accuracy.

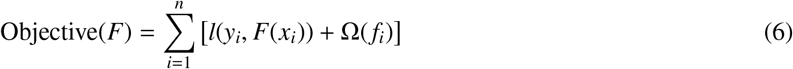

In this function, the initial portion accurately calculates the loss function (denoted as *l*(*y*_*i*_, *F*(*x*_*i*_))), effectively measuring the difference between the actual and predicted values of the output variable. Furthermore, the regularization penalty term (Ω(*f*_*i*_)) plays a crucial role in promoting a more simplified model. It is worth mentioning that this penalty term often incorporates a combination of L1 and L2 regularization techniques, as illustrated below:

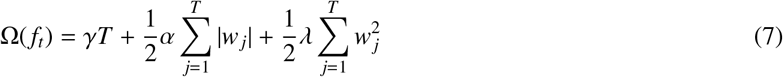

Within this mathematical expression, it is imperative to acknowledge the presence of three distinct regularisation parameters denoted as γ, α, and λ. The parameter γ, when multiplied by the number of leaf nodes represented by *T*, plays a pivotal role in regulating the complexity of the model. Moreover, both α and λ act as penalty coefficients pertaining to the L1 and L2 regularisation terms, respectively. These coefficients hold significant importance in attaining optimal regularisation outcomes, further amplifying the model’s simplicity and resilience.

The XGBoost algorithm possesses a remarkable advantage due to its capability to effectively manage a variety of data preprocessing tasks, which encompass activities like feature engineering, data normalization, and feature scaling. Consequently, this ability diminishes the necessity for extensive manual preprocessing, consequently saving valuable time and effort throughout the feature engineering phase as indicated by [30]. Furthermore, XGBoost integrates built-in functionalities designed to address missing values within the dataset, thus obviating the requirement for imputation techniques.

Simultaneously, XGBoost garners acclaim for its rapid processing speed and scalability, rendering it highly suitable for efficiently handling extensive datasets. It outshines numerous other machine learning algorithms in terms of both processing speed and predictive accuracy. Another notable attribute of XGBoost is its capacity to avert overfitting, thereby ensuring the generalization of unseen data instances. Nevertheless, XGBoost does confront certain constraints. One such limitation pertains to the substantial number of hyper-parameters it encompasses, which can pose challenges during the parameter tuning process, as highlighted in the study by [31]. Discovering the optimum combination of hyper-parameter values necessitates meticulous experimentation and can prove to be a time-intensive endeavour.

### 3.4. Proposed Hybrid Boosting Prediction Model

We devised an adaptive ensemble learning model to enhance the accuracy and reliability of predicting frequent ED visitors. The model incorporates an XGBoost algorithm as the foundation, augmented with a feature selection method that combines Elastic-Net and Local search techniques. To optimise the performance of XGBoost, we implemented a competitive approach using Differential Evolution (DE) and Nelder-Mead optimisation algorithms for hyper-parameter tuning. Besides, we evaluated three strategies for handling imbalanced data: under-sampling, oversampling (SMOTE), and adjusting class weights. By employing this comprehensive approach, we aimed to develop a robust prediction model that effectively identifies frequent ED visitors with improved precision and generalisability. Various components of the proposed prediction model are described as follows.

#### 3.4.1. Solutions for addressing imbalanced data

During our investigation, we encountered the challenge of imbalanced data, where the class distribution within the dataset exhibited a substantial disparity. This issue [32] arises when one class is represented by only a limited number of samples, referred to as the minority class. At the same time, the remaining samples belong to the other class, known as the majority class. Imbalanced data classification poses a predicament for classifiers as they tend to exhibit a bias towards the majority class, leading to imbalanced performance. This bias manifests in solutions favouring accuracy for the majority class but resulting in poor accuracy for the minority class. To address this issue, we explored and compared three techniques: over-sampling (employing synthetic samples through methods like SMOTE [33]), under-sampling [34] (randomly selecting a subset of samples from the majority class to achieve class balance while disregarding the remaining samples), and adjusting class weights [35] (assigning higher weights to samples from the minority class). Through our testing and analysis, we assessed the effectiveness of these techniques in mitigating the challenges posed by imbalanced data.

#### 3.4.2. Adaptive Elastic Net feature selection

With regard to selecting the most compelling features, we proposed a hybrid feature selection technique consisting of Elastic-net [36] combined with a fast local search called hill-climbing (HC) [37]. The role of HC is tuning the control parameters of Elastic-net during the feature selection to improve the performance of the most frequent visitor prediction model.

Elastic Net (as proposed by Zou and Hastie in 2005 [36]) is a sophisticated linear regression approach that cleverly marries the *L*1 regularisation technique of Lasso with the *L*2 regularisation method of Ridge regression. This unique fusion of regularisation methods allows Elastic Net to offer a balanced solution that benefits from the strengths of both Lasso and Ridge regression. The *L*1 regularization component in Elastic Net plays a crucial role in promoting sparsity within the solutions it generates, making it a valuable tool for feature selection as it can effectively drive numerous coefficients to zero. On the other hand, the *L*2 regularisation element in Elastic Net serves to combat overfitting by gently pulling the coefficients closer to zero, thus enhancing the model’s generalisation capabilities. Moving on to the scenario where *P* represents the count of predictors denoted by *x*_1_, *x*_2_, …, *x*_*P*_, the predicted value of the response variable *Y* can be expressed as *Y*^′^ = *ω*_0_ + *ω*_1_ *x*_1_ + *ω*_2_ *x*_2_ + … + *ω*_*P*_ *x*_*P*_ in line with the fundamental definition of linear regression. To determine the coefficients vector *ω*^′^ = [*ω*_0_, *ω*_1_, …, *ω*_*P*_]^*T*^, the objective is to minimise the sum of squared errors of residuals (SSE), which is represented as *S S E* = ||*Y*− *Xω*^′^||. In cases where the number of observations surpasses the dimension of features, the optimisation process involves minimising the loss function *L* to compute the coefficients in the following manner effectively:

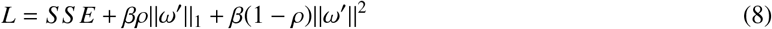

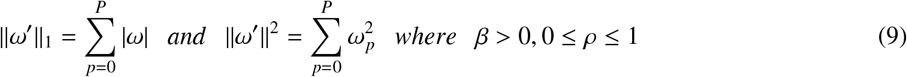

The penalty amount in Elastic Net is determined by adjusting weights and the parameter ρ. Fine-tuning both β and ρ is crucial for optimal results. Elastic Net has two special cases: ρ = 1 transforms it into LASSO, creating a sparse model with zero coefficients. ρ = 0 makes Elastic Net similar to ridge regression, allowing correlated predictors without restriction on the number of selected predictors.

Although Elastic Net’s capability shines in handling high-dimensional data, conducting feature selection, and managing correlated features [38], fine-tuning its control parameters presents a tough challenge. To address this issue, the current research introduces an efficient local search strategy designed to adjust the parameters of Elastic Net, encompassing L1-ratio, alpha, and selection methods (‘cyclic’, ‘random’). This optimisation technique adopts a heuristic search approach known as Hill climbing, which focuses on enhancing the Elastic Net’s configuration by incrementally refining it based on neighbouring solutions. The iterative nature of this method ensures that the Elastic Net gradually converges towards the most optimal configuration, striving to achieve the best possible solution. This iterative process persists until a local optimum is attained, signifying that no further enhancements can be achieved by transitioning to neighbouring solutions. The detailed steps of this adaptive Elastic Net approach are meticulously outlined in Algorithm 1, providing a comprehensive guide for implementing this sophisticated optimisation method.

#### 3.4.3. Hyper-parameters tuning

Tuning hyper-parameters in machine learning is of utmost importance due to its direct influence on the model’s performance and generalisation capabilities. The impact of hyper-parameter tuning is profound and multifaceted, as it directly affects how well a model can learn from data and make predictions. Indeed, this optimisation process aims to enhance the model’s accuracy, reduce error rates, and improve other relevant metrics, ultimately leading to better predictive performance. The ability to properly tune hyper-parameters enables the model to effectively capture the underlying patterns in the data, thereby enhancing its overall efficacy. Moreover, hyper-parameter tuning ensures that a machine learning model can generalise well to unseen data. Generalisation is critical to model performance, as it determines how well a model can predict new, unseen instances. Through hyper-parameter tuning, one can strike the right balance between under-fitting and overfitting.

In this study, we employed an effective optimisation method, Differential Evolution [39] (DE) and Nelder-mead [40] (NM), to fine-tune four hyper-parameters of the XGBoost model, which exhibited the best performance. The hyperparameters under consideration were the tree depth, number of estimators, learning rate, and sub-sample rate. We utilised both accuracy and AUC as metrics to adjust the XGBoost parameters effectively.

##### Algorithm 1

Adaptive Elastic Net feature selection

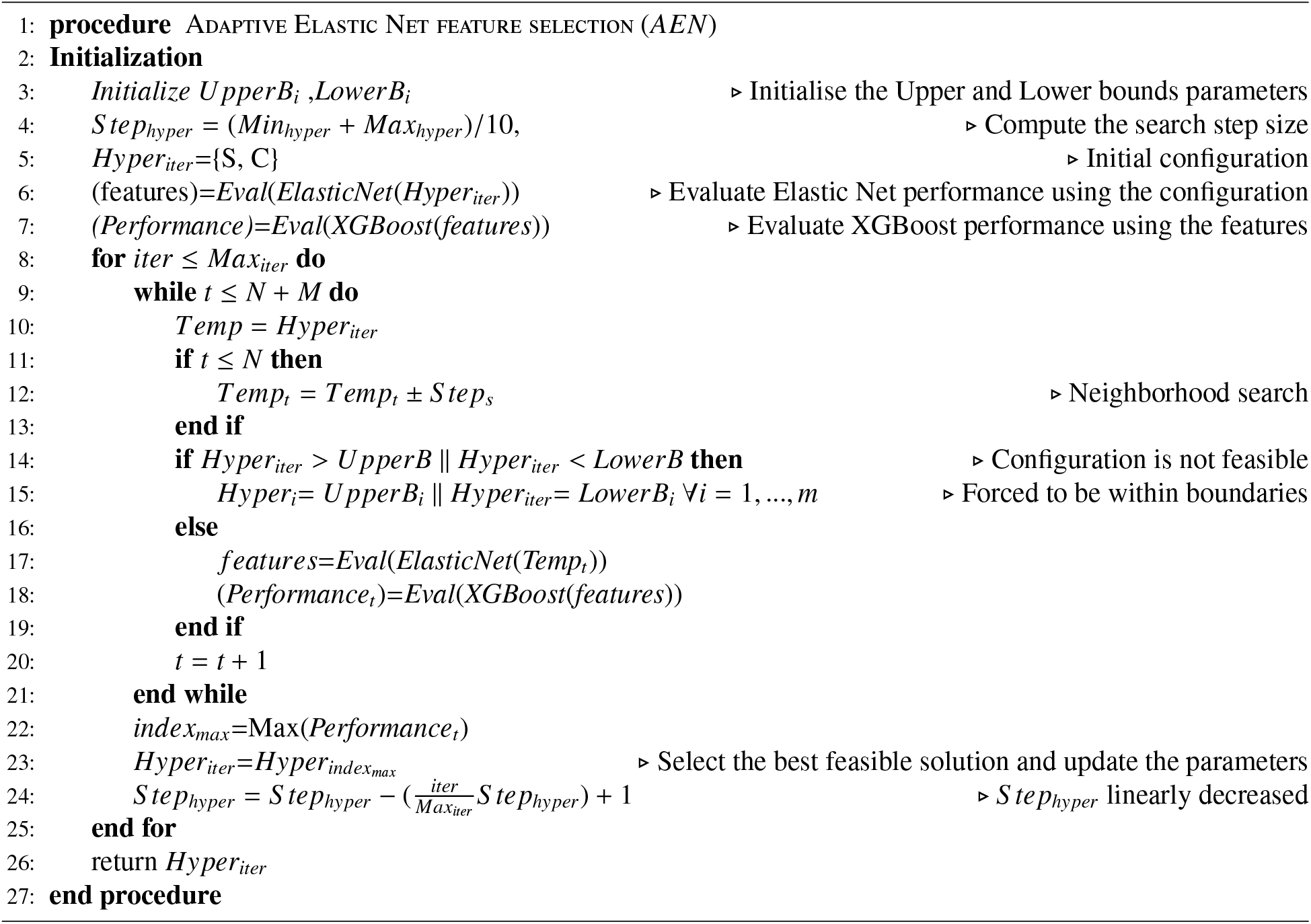

DE is a groundbreaking concept in evolutionary algorithms that incorporates differential vectors into a triangle search and has emerged as a highly favoured population-based optimisation technique, widely adopted for tackling a diverse array of real problems characterised by noise, dynamics, and multimodality. Among the various components of DE, the mutation operator stands out as a crucial element driving the algorithm’s success. Several mutation operators have been proposed within the realm of DE, each offering distinct convergence rates and exploration capabilities. One of the well-known mutation strategies is *DE*/*best*/1/*bin*, renowned for its ability to converge swiftly, particularly in handling unimodal problems. However, its efficacy diminishes when faced with challenges posed by local optima, often culminating in premature convergence while navigating through multi-modal problem landscapes. Equation (8) mathematically defines this mutation scheme, encapsulating its operational essence within DE.

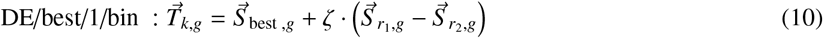

where 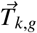 is a vector of differential of three solutions, including 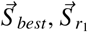, and 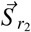. In the meantime, ζ is the mutation factor which can adjust the step size and speed of exploration ability. Moving on to another evolutionary operator within DE, we encounter the pivotal role of crossover. The predominant form of crossover method employed is the binomial approach, characterised by a formulation that involves the trial vector, denoted as 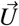, and the crossover rate, symbolised as *C*_*r*_, ranging from zero to one as follows.

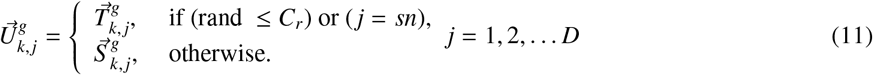

During the crossover process, a specific number of candidates, represented by *sn*, are selected. This culminates in the creation of a novel solution through a reasonable combination of the parent and offspring elements, perpetuating the evolutionary cycle of optimisation as follows.

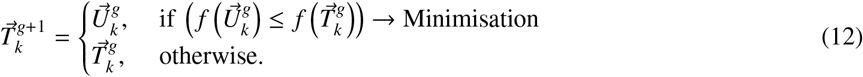

## 4. Experimental results and discussions

In this section, to identify the most effective classification model for predicting frequent ED visitors, we conducted a comparative analysis of 16 popular machine-learning algorithms. These models encompassed logistic regression (LR), Stochastic gradient descent (SGD), k-nearest neighbours (KNN), three neural networks (MLP, Perceptron, and DNN), Decision trees (DT), Extra trees [41] (ET), Random forests [42] (RF), and six ensemble boosting models (XGBoost [29], AdaBoost [43], CatBoost [44], GBM [28], LightGBM [45], and HGBM [46]).

The prediction results of these models are detailed in Section 4.1. Subsequently, we explored the impact of two optimisation techniques, DE and Nelder-Mead, on tuning the hyper-parameters of the best-performing classifier, XG-Boost. The landscape analysis and AUC convergence results are provided in Section 4.3. Furthermore, we extensively discussed the performance of the proposed feature selection method in Section 4.4. Lastly, we presented a comparative evaluation of three approaches for addressing imbalanced data in Section 4.1. These comprehensive analyses and comparisons shed light on the most accurate and robust techniques for predicting frequent ED visitors, contributing to developing an effective classification model.

### 4.1 Fundamental prediction results

The performance of nine ensemble learning models and seven ML methods was assessed, and the statistical results are presented in Figure 6. Due to utilising 10-fold cross-validation for classifier validation, each box plot represents the minimum, maximum, and median metrics, with outliers indicated by a plus sign. Analysis of Figure 6(a), (b), (c), and (d) reveals that XGBoost consistently outperforms other models, except for precision, where LightGBM and GBM exhibit superior performance. However, all nine ensemble models demonstrate competitive performance, with accuracy ranging from 76.6% to 78.2%. On average, ensemble models outperform other ML models, including LR, NNs, and KNN. Supplementary Table A.4 provides the formulation of evaluation metrics employed in this study. Table 5 and Supplementary Table A.5 present detailed classification results for eight ensemble and eight ML models, respectively, with six evaluation metrics. Regarding accuracy, XGBoost is the top-performing model, with an average of 78%. Notably, LightGBM demonstrates high precision at 85.7% and outperforms other models in this regard.

**Table 5:** Statistical results of eight ensemble learning models with six performance evaluation metrics for predicting the frequent ED visitors (¿=3 visits per year)

|  | AdaBoost |  |  |  |  |  | CatBoost |  |  |  |  |  |  |
| --- | --- | --- | --- | --- | --- | --- | --- | --- | --- | --- | --- | --- | --- |
|  | Accuracy | Precision | Recall | F1_score | AUC | log_loss |  | Accuracy | Precision | Recall | F1_score | AUC | log_loss |
| Min | 0.766 | 0.556 | 0.003 | 0.007 | 0.501 | 7.983 | Min | 0.773 | 0.633 | 0.065 | 0.118 | 0.527 | 7.730 |
| Max | 0.769 | 0.661 | 0.004 | 0.008 | 0.502 | 8.071 | Max | 0.776 | 0.653 | 0.071 | 0.128 | 0.529 | 7.838 |
| Mean | 0.768 | 0.585 | 0.004 | 0.008 | 0.502 | 8.024 | Mean | 0.774 | 0.642 | 0.068 | 0.123 | 0.528 | 7.790 |
| Median | 0.768 | 0.573 | 0.004 | 0.008 | 0.502 | 8.029 | Median | 0.775 | 0.640 | 0.069 | 0.124 | 0.528 | 7.786 |
| STD | 8.76E-04 | 3.67E-02 | 3.09E-04 | 6.11E-04 | 1.52E-04 | 3.02E-02 | STD | 1.04E-03 | 6.66E-03 | 2.03E-03 | 3.36E-03 | 9.21E-04 | 3.61E-02 |
|  | XGBoost |  |  |  |  |  | HGBM |  |  |  |  |  |  |
|  | Accuracy | Precision | Recall | F1_score | AUC | log_loss |  | Accuracy | Precision | Recall | F1_score | AUC | log_loss |
| Min | 0.779 | 0.620 | 0.131 | 0.216 | 0.553 | 7.534 | Min | 0.770 | 0.658 | 0.030 | 0.058 | 0.513 | 7.848 |
| Max | 0.782 | 0.638 | 0.139 | 0.228 | 0.557 | 7.632 | Max | 0.773 | 0.674 | 0.036 | 0.069 | 0.515 | 7.947 |
| Mean | 0.780 | 0.630 | 0.134 | 0.221 | 0.555 | 7.586 | Mean | 0.771 | 0.666 | 0.034 | 0.064 | 0.514 | 7.898 |
| Median | 0.780 | 0.630 | 0.135 | 0.222 | 0.556 | 7.594 | Median | 0.771 | 0.666 | 0.034 | 0.064 | 0.514 | 7.902 |
| STD | 1.02E-03 | 5.34E-03 | 3.03E-03 | 4.37E-03 | 1.46E-03 | 3.54E-02 | STD | 1.06E-03 | 5.03E-03 | 1.82E-03 | 3.31E-03 | 7.95E-04 | 3.65E-02 |
|  | LightGBM |  |  |  |  |  | Extra Tree |  |  |  |  |  |  |
|  | Accuracy | Precision | Recall | F1_score | AUC | log_loss |  | Accuracy | Precision | Recall | F1_score | AUC | log_loss |
| Min | 0.766 | 0.814 | 0.000 | 0.001 | 0.500 | 7.985 | Min | 0.767 | 0.688 | 0.011 | 0.021 | 0.505 | 7.934 |
| Max | 0.769 | 0.917 | 0.001 | 0.003 | 0.501 | 8.072 | Max | 0.770 | 0.739 | 0.012 | 0.025 | 0.505 | 8.031 |
| Mean | 0.768 | 0.857 | 0.001 | 0.002 | 0.500 | 8.026 | Mean | 0.769 | 0.705 | 0.012 | 0.023 | 0.505 | 7.979 |
| Median | 0.768 | 0.845 | 0.001 | 0.002 | 0.501 | 8.029 | Median | 0.769 | 0.703 | 0.011 | 0.022 | 0.505 | 7.981 |
| STD | 9.37E-04 | 3.63E-02 | 3.09E-04 | 6.18E-04 | 1.48E-04 | 3.23E-02 | STD | 9.52E-04 | 1.59E-02 | 5.89E-04 | 1.14E-03 | 2.58E-04 | 3.29E-02 |
|  | Random Forest |  |  |  |  |  | Decssion Tree |  |  |  |  |  |  |
|  | Accuracy | Precision | Recall | F1_score | AUC | log_loss |  | Accuracy | Precision | Recall | F1_score | AUC | log_loss |
| Min | 0.769 | 0.738 | 0.020 | 0.039 | 0.509 | 7.876 | Min | 0.767 | 0.537 | 0.027 | 0.052 | 0.510 | 7.944 |
| Max | 0.772 | 0.791 | 0.022 | 0.043 | 0.510 | 7.969 | Max | 0.770 | 0.571 | 0.035 | 0.066 | 0.513 | 8.036 |
| Mean | 0.771 | 0.756 | 0.021 | 0.041 | 0.509 | 7.918 | Mean | 0.769 | 0.551 | 0.030 | 0.057 | 0.511 | 7.988 |
| Median | 0.771 | 0.747 | 0.021 | 0.041 | 0.509 | 7.918 | Median | 0.769 | 0.551 | 0.029 | 0.055 | 0.511 | 7.991 |
| STD | 8.98E-04 | 1.78E-02 | 7.85E-04 | 1.49E-03 | 3.91E-04 | 3.10E-02 | STD | 8.78E-04 | 1.02E-02 | 2.76E-03 | 4.93E-03 | 9.90E-04 | 3.03E-02 |

**Figure 6:**
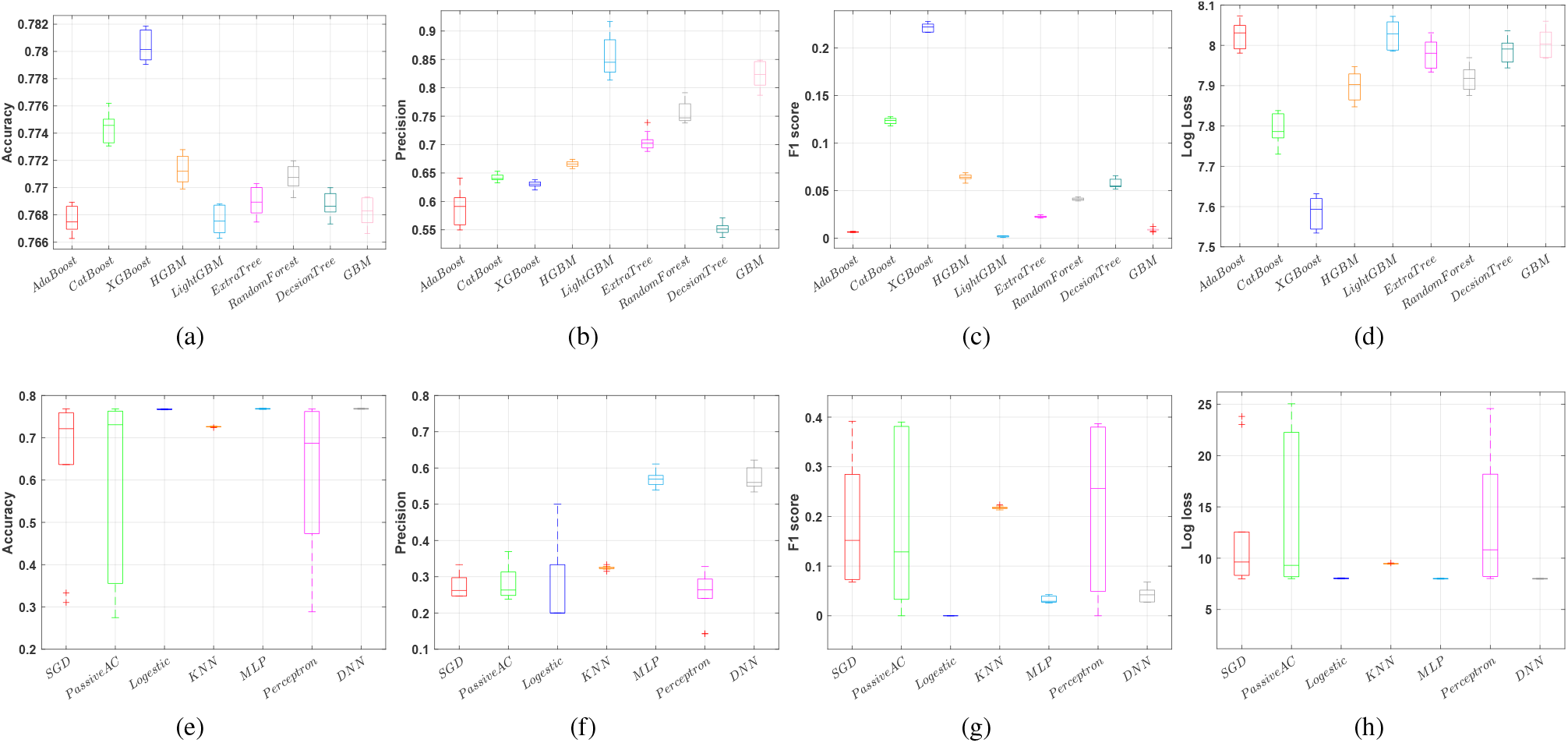
Statistical results of nine ensemble learning models and seven popular ML methods in predicting the frequent ED visitor (+3). 10-fold cross-validation was used to evaluate the performance of the models. For accuracy, precision and F1-score, higher values indicate better performance; however, a low value for Log loss shows that the model is performing well in terms of its predictive accuracy.

We employed the Friedman test, a ranked-based statistical test, to determine the best-performing classifier among the 16 models utilised. This test extends the Wilcoxon signed-rank test and is the nonparametric equivalent of one-way repeated measures. We calculated the rank of each model based on all evaluation metrics and presented the results of the Friedman test in Table 6. Particularly, XGBoost achieved the highest rank compared to other models in terms of accuracy, F1 score, and log loss. While our models have demonstrated high accuracy and precision, the AUC metric presents an area for improvement, as noted in Table 5. The AUC reflects the trade-off between the true positive rate and the false positive rate, and achieving a higher AUC value is desirable. To address this concern and enhance the AUC performance, we have explored three methods, which are discussed in the subsequent section. These methods aim to rectify the issue and optimise the balance between true positive and false positive rates, ultimately improving the AUC metric.

**Table 6:** Mean ranks and *p* − *value* computed by Friedman test for 16 machine learning models with four performance evaluation metrics for predicting the frequent ED visitors (¿=3 visits per year)

| Metric | XGBoost | AdaBoost | CatBoost | HGBM | LightGBM | Extra Tree | Random Forest | Decision Tree |
| --- | --- | --- | --- | --- | --- | --- | --- | --- |
| Accuracy | **1.0** | 10.5 | 2.0 | 3.0 | 10.5 | 5.1 | 4.0 | 6.4 |
| Precision | 7.1 | 8.8 | 6.3 | 5.1 | **1.4** | 4.1 | 3.1 | 10.5 |
| F1 Score | **3.0** | 13.6 | 4.7 | 6.6 | 14.7 | 11.6 | 9.1 | 7.5 |
| Log loss | **1.0** | 10.5 | 2.0 | 3.0 | 10.5 | 5.1 | 4.0 | 6.4 |
| Average Rank | **3.0** | 10.9 | 3.8 | 4.4 | 9.3 | 6.5 | 5.1 | 7.7 |
| P-value | 0.0E+00 | 1.1E-39 | 7.8E-28 | 8.1E-35 | 9.9E-39 | 1.8E-37 | 2.6E-35 | 6.6E-40 |
| Metric | GBM | SGD | Passive AC | Logestic | KNN | MLP | Perceptron | DNN |
| Accuracy | 8.6 | 14.1 | 14.6 | 12.3 | 14.3 | 8.0 | 14.7 | 7.0 |
| Precision | 1.8 | 14.0 | 14.2 | 13.4 | 12.8 | 9.5 | 14.5 | 9.4 |
| F1 Score | 12.6 | 3.6 | 5.7 | 15.7 | 3.2 | 10.3 | 5.2 | 8.9 |
| Log loss | 8.5 | 14.1 | 14.6 | 12.3 | 14.3 | 8.0 | 14.7 | 7.0 |
| Average Rank | 7.9 | 11.5 | 12.3 | 13.4 | 11.2 | 8.9 | 12.3 | 8.1 |
| P-value | 1.8E-36 | 1.5E-04 | 9.5E-05 | 1.7E-39 | 1.4E-48 | 1.7E-38 | 5.2E-06 | 7.8E-40 |

### 4.2. The results of handling imbalanced data

To address the challenge of imbalanced data and improve the classifier’s AUC performance, we conducted comparative experiments using three methods: over-sampling (SMOTE), under-sampling, and adjusting class weights. Initially, we identified the best-performing model among the 16 ML models: XGBoost. Subsequently, we examined the impact of different weights assigned to the minority (positive) class, ranging from two to five. Figure 7 illustrates the performance of XGBoost with weighted class in terms of AUC and accuracy. Interestingly, we observed a direct relationship between increasing the class weight and AUC. However, there was a decrease in accuracy when raising the class weight from 80% to 67%. Therefore, it is crucial to identify an optimal weight that strikes a balance between AUC and accuracy. In this particular case study, the optimal weight was determined to be 3, representing a suitable trade-off between AUC performance and accuracy.

**Figure 7:**
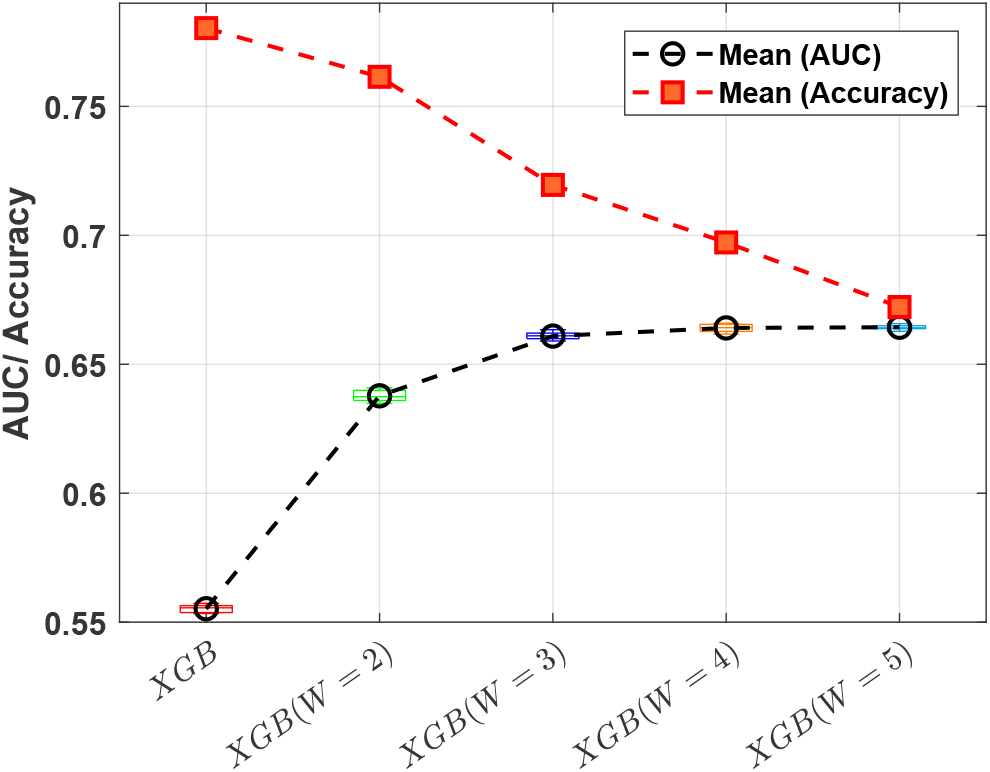
A trade-off between the Accuracy and AUC using adjusted class weight for XGBoost.

Figure 8 presents a comparison of three imbalanced data handling techniques, along with the original XGBoost model, using four evaluation metrics. When considering AUC, the methods of adjusting class weight and undersampling emerge as the best performers, achieving rates of 68% and 67%, respectively. Notably, these two methods have substantially improved the recall rate from 21% to 55% and 68%. Enhancing recall is of paramount importance as it enables the model to more accurately identify positive instances, thereby reducing the occurrence of false negatives. This is particularly significant in domains where the repercussions of missing positive instances, such as in medical diagnoses or fraud detection, can be severe. A higher recall rate signifies a reduced number of missed positive instances, resulting in heightened sensitivity and a more reliable identification of true positives.

**Figure 8:**
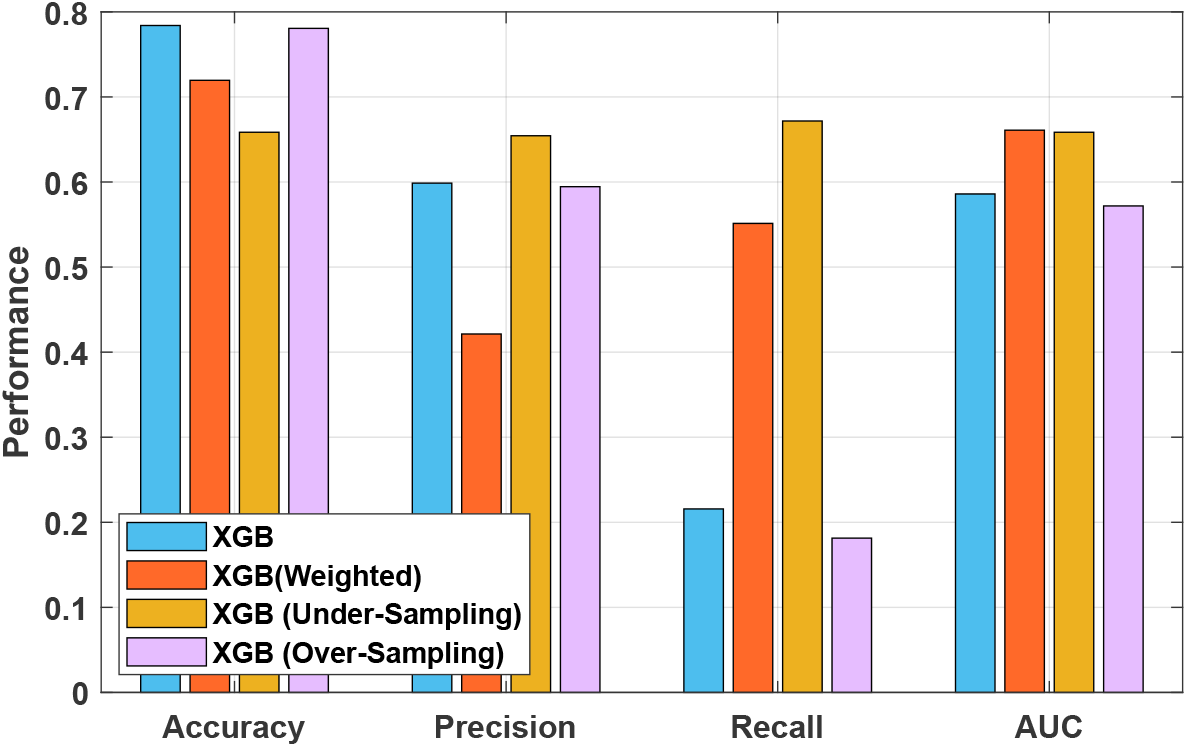
Compared techniques to mitigate the problem of imbalanced data using XGBoost in predicting the frequent ED visitors (+3).

### 4.3. Hyper-parameters tuning

Figure 9 illustrates the landscape of hyper-parameter tuning for XGBoost. The results indicate that increasing the depth of trees enhances accuracy; however, for achieving a high AUC, the optimal range appears to be around 10. Moreover, we observed that a larger number of estimators yielded better performance. Interestingly, utilising the entire training dataset (subsample=1.0) for training the trees demonstrated the highest accuracy and AUC. Finally, based on Figure 9 (d), the optimal learning rate value was found to be approximately 0.4.

**Figure 9:**
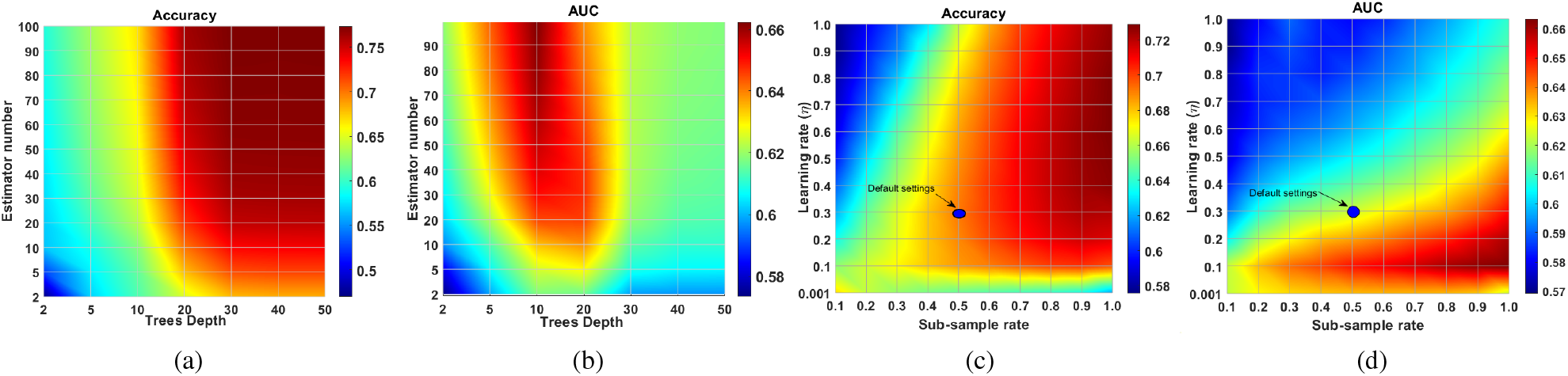
The landscape of Hyper-parameters tuning using Grid search for a number of estimators, depth of trees, sub-sample rate and learning rate. Higher values indicate better performance for accuracy and AUC.

In order to demonstrate the convergence speed of both DE and Nelder-Mead in optimising the hyper-parameters of XGBoost, we conducted ten independent runs for each optimiser with randomly initialised parameters. Figure 10 indicates the convergence pattern of the two optimisers. Nelder-Mead exhibits rapid initial convergence within the first few evaluations (less than 200); however, it becomes trapped in a local optimum and fails to improve the AUC further. Conversely, although DE has a slower convergence rate compared to Nelder-Mead, it maintains an upward trend throughout the optimisation process and ultimately surpasses Nelder-Mead’s performance. Therefore, if the training runtime is not a constraint, we recommend utilising DE for hyper-parameters tuning as it demonstrates better overall performance. Conversely, if runtime is a crucial factor, Nelder-Mead can be a more suitable choice for fine-tuning the hyper-parameters.

**Figure 10:**
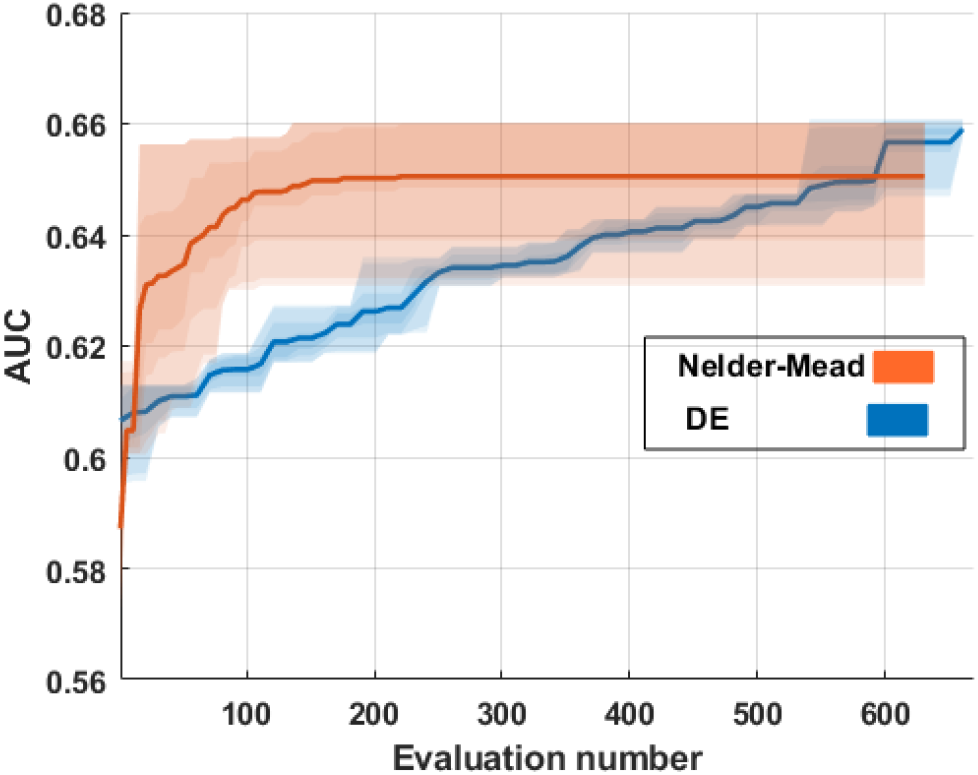
Hyper-parameters tuning using DE and Nelder-mead based on AUC.

### 4.4. Feature selection results

The effectiveness of the proposed adaptive method for optimal feature selection, as outlined in Section 3.4.2, is demonstrated in Figure 11. The graph showcases the efficiency achieved when utilising the optimal subset of features. It is observed that increasing the number of optimal features from five to seven leads to improvements in both accuracy and AUC, with gains of approximately 6% and 4%, respectively. However, beyond seven features, both accuracy and AUC experienced a decline. Consequently, the optimal number of features for this particular scenario is seven. These seven features comprise Age, Sex, ICD10, Disposition, Year (arrival time in ED), State, and Suburb, which collectively offer the best performance in terms of accuracy and AUC for the given dataset and classification task.

**Figure 11:**
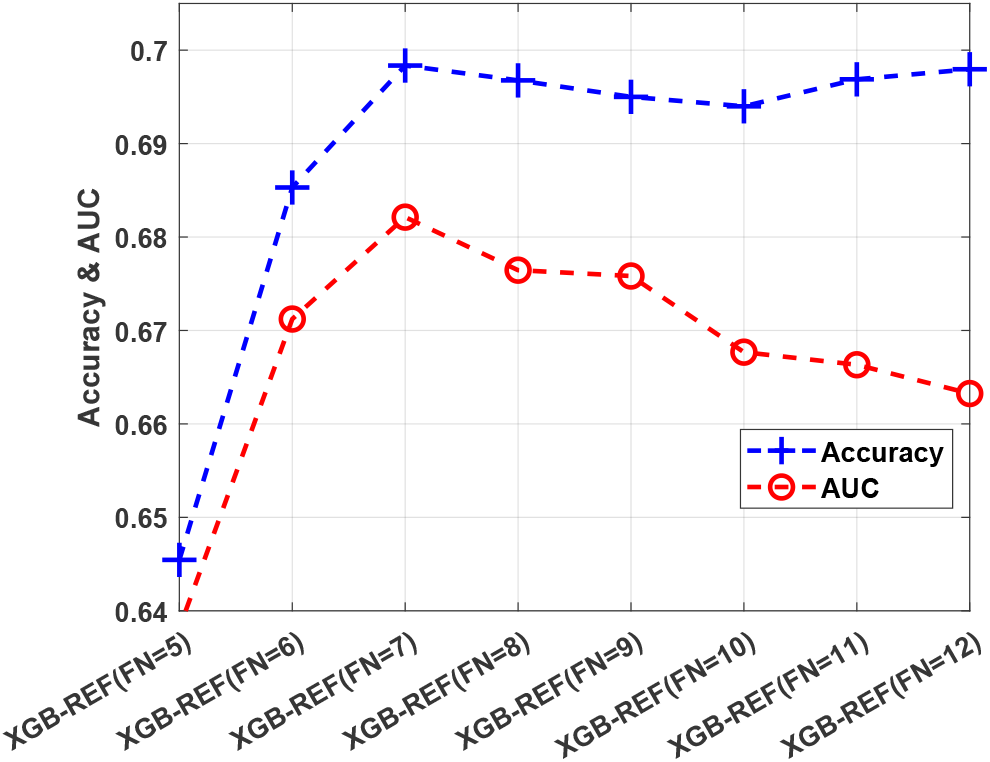
Feature selection impact on the performance of XGBoost in predicting the frequent ED visitors (+3). The best feature number and configuration is seven, including Age, Sex, ICD10, Disposition, Year (arrival time in ED), State, and Suburb.

## 5. Conclusions

Frequent ED visitors are a significant concern within primary and emergency care settings. Machine learning models have gained popularity in medicine and healthcare and are being increasingly utilised. ML models continuously advance, present novel possibilities, and witness notable theoretical advancements. These advancements have led to improved accuracy and enhanced reliability of ML models.

In this research, an Adaptive ensemble learning–based prediction model was devised and evaluated against 16 prominent machine learning models using a large-scale dataset collected from Canberra Hospital, a tertiary public hospital in ACT, Australia, between 1997 and 2022 with 1.6 million records of patients’ episodes. Also, three ways of adjusting were examined to deal with the challenge of imbalanced data. At the same time, a combination of two feature selection approaches incorporating Elastic-Net and local search was proposed to identify the best feature combination. Hyper-parameter tuning was managed and done using a population-based set of computer instructions and a local search. The performance of these ways of doing things was compared to determine their effectiveness in optimising the model’s abilities.

The experimental modelling results revealed that the proposed model considerably outperformed other models in terms of five metrics: accuracy, Recall, F1-score, Area under the ROC curve (AUC), and Log loss at 0.78 (95% CI 0.78-0.79), 0.68 (95% CI 0.68-0.68), 0.68 (95% CI 0.68-0.69), 0.68 (95% CI 0.69-0.70), and 7.4 (95% CI 7.2-7.5), respectively.

Our future research aims to explore the potential impact of additional features on enhancing prediction results. Specifically, we plan to investigate the inclusion of weather parameters and pollen count levels, as these factors have been found to influence various health conditions. Furthermore, we aim to expand our analysis to encompass a higher rate of ED visits, such as ten visits per year, to gain a deeper understanding of the predictive capabilities of our model. Additionally, we intend to explore time-series forecasting machine learning models as part of our methodology. By incorporating these advancements, we anticipate further improving the accuracy and reliability of our predictions in the context of frequent ED visits.

## Data Availability

All data produced in the present study are available upon reasonable request to Michael Phipps, the esteemed Senior Director of the Canberra Health Service.

## Ethics approval and consent to participate

### Data and Code availability

For information regarding the availability of data used in this study, interested individuals can obtain access by contacting the corresponding author, Michael Phipps. Python 3.9.16 served as the programming language for implementing this study. Various open-source Python libraries were utilized, including Scikit-learn (0.23.2) ^1^, NumPy (1.19.1), Pandas (1.1.0), SciPy (1.5.2), and TensorFlow (2.3.0). Scikit-learn played a central role in supplying a wide range of machine learning models employed in this research. Furthermore, ensemble models such as XGBoost ^2^, LightGBM ^3^, and CatBoost^4^, accessible open-source platforms were utilized. TensorFlow ^5^, an open-source machine learning framework developed by Google, was specifically employed for implementing deep neural networks (DNN) and convolutional deep neural networks (CDNN). In order to tune the hyper-parameters, we applied a well-known optimisation method, Differential Evolution (DE)^6^, from the Scipy ^7^ library. By combining these libraries and frameworks, a comprehensive array of tools and algorithms were utilized to develop and analyze machine learning models effectively.

## Declaration of conflicting interests

The authors declared no potential conflicts of interest with respect to the research, authorship, and/or publication of this article.

## Ethics approval

This study has received ethical approval from The ACT Health Human Research Ethics Committee (Approval No. 2024/ETH01037).

## Data availability

The dataset regarding ED Presentations used in this research is owned by Canberra Health Services and the ACT Health Directorate. Access to data for our study relies on obtaining anonymised datasets, a task made possible through collaboration with Michael Phipps, the esteemed Senior Director of the Canberra Health Service.

## Appendix A. Supplementary materials

**Table A.1:** Descriptions of the features considered in this study.

| Num | Features name | Description |
| --- | --- | --- |
| 1 | MRN | Patient ID |
| 2 | Episode | Unique ID of the episode |
| 3 | Treat-Doc-Date | The time a junior doctor sees a patient |
| 4 | SNR-Doc-Date | The time a senior doctor sees a patient |
| 5 | Dep-Ready-Date | The time a patient is ready to depart. For patients who are admitted, this is the end of clinical care and the start of bed block. For patients going home, this is the time that clinical care ends |
| 6 | Admit-Date | For admitted patients, the time that they are admitted |
| 7 | Sex | Female or male |
| 8 | Age | Age at episode commencement |
| 9 | Suburb | suburb |
| 10 | State | State (NSW/ACT etc.) |
| 11 | Diag-ICD | The ICD9 and ICD10 code |
| 12 | Diag-Desc | Description of the ICD9 and 10 code |
| 13 | DS-Desc | the disposition of the patient (Admit / Did Not Wait (DNW) / Left Own Risk (LOR), Home, transferred to another hospital, died |
| 14 | DS-Extract-Code | Non-admitted patient emergency department service episode—episode end status, code N (https://meteor.aihw.gov.au/content/746709) |
| 15 | UN-Code | Ward the patient was admitted to (if admitted) |
| 16 | SP-Code | The speciality a patient was admitted to |
| 17 | Earliest DTTM | The arrival time for the patient in ED |
| 18 | Triage Priority | Triage category |

**Table A.4:** The performance evaluation metrics.

| Abbreviation | full name | formula |
| --- | --- | --- |
| TPR | True Positive Rate | $= \frac{TP}{TP+FN}$ |
| FPR | False Positive Rate | $= \frac{FP}{TN+FP}$ |
| PPV | Positive Predictive Value | $= \frac{TP}{TP+FP}$ |
| ACC | Accuracy | $= \frac{TP+TN}{TP+TN+FP+FN}$ |
| PRE | Precision | $= \frac{TP}{TP+FP}$ |
| REC | Recall | $= \frac{TP}{TP+FN}$ |
| SPF | Specificity | $= \frac{TN}{TN+FP}$ |
| AUC | Area Under the ROC Curve | $= \int_0^1 TPR(FPR^{-1}(t)), dt$ |
| F1-score | Harmonic mean of PPV and recall | $= \frac{2 \cdot PPV \cdot Recall}{PPV + Recall} = \frac{2 \cdot TP}{2 \cdot TP + FP + FN}$ |

**Table A.2:** The technical review of the Deep and Machine learning methods in predicting frequent ED visitors.

| Ref | Title | Dataset features | Case study | Date range | Sample size | AI models | Best-performed method | Accuracy |
| --- | --- | --- | --- | --- | --- | --- | --- | --- |
| [2] | ML to improve frequent ED use prediction: a retrospective cohort study | Demographic: sex, age, birth and death date, patient residence place; Physician reimbursement: service date, place of service place (emergency, medical clinic, etc.), physician speciality, diagnosis (ICD-9). A medical act done by the physician: The hospital register, which contains information about the reasons for hospitalisation (main diagnosis and up to 25 secondary diagnoses coded in ICD-10), dates of admission and release from the hospital, and all medical procedures performed during the hospitalisation. | Quebec (Canada), age≥18 | Jan 2012 until the Dec 2013, ED visit frequency≥4 | 43K and 15K | Gradient boosting machines (GBM), Naïve Bayes (NB), Neural networks (NN), Random forests (RF) | GBM | AUC=81.4, SEN=69.9, SPE=78.9, PPV=9.3, NPV=98.8 |
| [5] | Frequent ED use by older adults with ambulatory care sensitive conditions | age, presence of chronic obstructive pulmonary disorder or diabetes, higher comorbidity index, common mental health disorders, polypharmacy, higher number of past ED and specialist visits, rural residence, and higher material and social deprivation | Quebec (Canada) age≥65 years | 2012 and 2013, ED visit frequency≥4 | 17K | Multivariate logistic regression | Focused on one method | reported odd ratios (OR) with the associated 99% confidence intervals (CI). |
| [47] | Comparative analysis of ML approaches for predicting frequent ED visits | demographics (i.e. age, gender, type of insurance, residential region); (ii) reason for visit; (iii) access to ED (i.e. season of visit, day of the visit, type of transportation visited ED (private vs public); and (iv) hospitalization (i.e. admitted/discharged after initial ED visit). | Korea Institute for Health and Social Affairs | between 2008 and 2015, ED visit frequency ≥4 | 9K | Logistic regression, SVM, Random forest, Bagging, and Voting | Random forest | Classification error= 3.77, % Sensitivity= 0.95, PPV= 0.98, AUC= 0.96, Calibration error= 10.3. % |
| [18] | Risk Prediction of ED Visits in Patients With Lung Cancer Using ML | Demographics, Visit history, Clinical information, Laboratory Test, Vital Signs | Seoul National University Bundang Hospital, age≥18 | Jan 2010 and Dec 2017 | 7K | logistic regression, random forest, XGB, and light gradient boosting machine (LGBM) | LGBM | Sensitivity=0.76, Specificity= 0.58, AU-ROC= 0.73, PRAUC= 0.24 |
| [48] | Predicting frequent ED use by people with epilepsy | Demographics (age, 3-digit zip code), visit history (ED, inpatient, and outpatient), radiology encounters, and comorbidities (ICD-9 codes collapsed into 33 comorbidities) | NYCLIX (New York Clinical Information Exchange) | 2009 to 2012 | 8K | Logistic regression, Lasso, Elastic net, CART, Random Forests, AdaBoost, SVM | AdaBoost | Classification error= 5.32, AUC= 0.88 Sensitivity= 30, PPV=63, Calibration=5 |
| [49] | A ML approach to predicting need for hospitalization for pediatric asthma exacerbation at the time of ED triage | demographic information (including patient sex, age, race, and ethnicity), patient acuity at triage (measured by the Emergency Severity Index [ESI]), and the first recorded measurement of (weight, oxygen saturation, heart rate, and respiratory rate). | children's hospital UK(2 ≤ age ≤ 18) | Jan 1, 2012, and Dec 31, 2015 | 29K | Decision trees, LASSO, random forests, and GBM | GBM | AUC= 84 |

**Table A.3:** Multivariable analysis to examine the association between frequent ED visits and various age groups, including odds ratio (OR) with 95% confidence intervals (CIs). The analysis included 253K female and 276K male patients, with a total of 787K and 822K visits, respectively, spanning from 1997 to 2022. The results revealed a significant association between frequent ED visits and male patients over the age of 64, with an OR identified at 1.02. The statistical significance of the interaction between frequent ED visitors (3+ visits per year) and age was established across all age groups, as indicated by the p-value being less than 0.05.

| Covariate | Odds Ratio | CI Lower | CI Upper | P-value |
| --- | --- | --- | --- | --- |
| Sex (female) * ($age < 14$) | 0.485 | 0.480 | 0.490 | 0 |
| Sex (female) * ($14 \leq age < 24$) | 0.869 | 0.860 | 0.879 | 1.20E-135 |
| Sex (female) * ($24 \leq age < 48$) | 0.733 | 0.727 | 0.739 | 0 |
| Sex (female) * ($48 \leq age < 64$) | 0.607 | 0.600 | 0.615 | 0 |
| Sex (female) * ($age \geq 64$) | 0.886 | 0.877 | 0.895 | 1.39E-114 |
| Sex (male) * ($age < 14$) | 0.485 | 0.481 | 0.490 | 0 |
| Sex (male) * ($14 \leq age < 24$) | 0.508 | 0.502 | 0.514 | 0 |
| Sex (male) * ($24 \leq age < 48$) | 0.583 | 0.578 | 0.588 | 0 |
| Sex (male) * ($48 \leq age < 64$) | 0.624 | 0.617 | 0.632 | 0 |
| Sex (male) * ($age \geq 64$) | **1.022** | 1.011 | 1.033 | 8.46E-05 |

**Table A.5:** Statistical results of eight machine learning models with six performance evaluation metrics for predicting the frequent ED visitors (¿=3 visits per year)

|  | MLP |  |  |  |  |  | DNN |  |  |  |  |  |  |
| --- | --- | --- | --- | --- | --- | --- | --- | --- | --- | --- | --- | --- | --- |
|  | Accuracy | Precision | Recall | F1_score | AUC | log_loss |  | Accuracy | Precision | Recall | F1_score | AUC | log_loss |
| Min | 0.767 | 0.539 | 0.013 | 0.026 | 0.505 | 7.953 | Min | 0.767 | 0.534 | 0.014 | 0.027 | 0.506 | 7.947 |
| Max | 0.770 | 0.611 | 0.022 | 0.043 | 0.509 | 8.048 | Max | 0.770 | 0.622 | 0.036 | 0.068 | 0.513 | 8.035 |
| Mean | 0.768 | 0.570 | 0.017 | 0.033 | 0.507 | 8.000 | Mean | 0.769 | 0.572 | 0.022 | 0.042 | 0.508 | 7.993 |
| Median | 0.768 | 0.569 | 0.015 | 0.029 | 0.506 | 8.003 | Median | 0.768 | 0.560 | 0.022 | 0.042 | 0.508 | 8.001 |
| STD | 8.78E-04 | 2.11E-02 | 3.57E-03 | 6.70E-03 | 1.32E-03 | 3.03E-02 | STD | 9.21E-04 | 3.11E-02 | 7.60E-03 | 1.39E-02 | 2.64E-03 | 3.18E-02 |
|  | SGD |  |  |  |  |  | Perceptron NN |  |  |  |  |  |  |
|  | Accuracy | Precision | Recall | F1_score | AUC | log_loss |  | Accuracy | Precision | Recall | F1_score | AUC | log_loss |
| Min | 0.311 | 0.000 | 0.000 | 0.000 | 0.500 | 7.996 | Min | 0.289 | 0.000 | 0.000 | 0.000 | 0.500 | 8.020 |
| Max | 0.768 | 0.333 | 0.951 | 0.392 | 0.538 | 23.813 | Max | 0.768 | 0.328 | 0.945 | 0.387 | 0.548 | 24.573 |
| Mean | 0.645 | 0.222 | 0.275 | 0.178 | 0.516 | 12.255 | Mean | 0.605 | 0.236 | 0.369 | 0.230 | 0.523 | 13.645 |
| Median | 0.721 | 0.262 | 0.119 | 0.152 | 0.512 | 9.623 | Median | 0.687 | 0.264 | 0.232 | 0.256 | 0.527 | 10.815 |
| STD | 1.76E-01 | 1.20E-01 | 3.62E-01 | 1.52E-01 | 1.47E-02 | 6.08E+00 | STD | 1.79E-01 | 9.65E-02 | 3.61E-01 | 1.57E-01 | 1.67E-02 | 6.20E+00 |
|  | Passive Aggressive Classifier |  |  |  |  |  | Logistic Classifier |  |  |  |  |  |  |
|  | Accuracy | Precision | Recall | F1_score | AUC | log_loss |  | Accuracy | Precision | Recall | F1_score | AUC | log_loss |
| Min | 0.275 | 0.238 | 0.000 | 0.000 | 0.500 | 8.006 | Min | 0.766 | 0.000 | 0.00E+00 | 0.00E+00 | 0.500 | 7.989 |
| Max | 0.768 | 0.500 | 0.959 | 0.390 | 0.538 | 25.047 | Max | 0.769 | 1.000 | 2.68E-05 | 5.36E-05 | 0.500 | 8.080 |
| Mean | 0.617 | 0.307 | 0.324 | 0.181 | 0.515 | 13.230 | Mean | 0.767 | 0.237 | 1.33E-05 | 2.67E-05 | 0.500 | 8.033 |
| Median | 0.731 | 0.278 | 0.087 | 0.129 | 0.509 | 9.300 | Median | 0.767 | 0.100 | 1.33E-05 | 2.65E-05 | 0.500 | 8.037 |
| STD | 2.05E-01 | 8.02E-02 | 4.15E-01 | 1.62E-01 | 1.51E-02 | 7.09E+00 | STD | 9.29E-04 | 3.25E-01 | 1.41E-05 | 2.81E-05 | 9.88E-06 | 3.21E-02 |
|  | KNN |  |  |  |  |  | Extreme Learning Machine (ELM) |  |  |  |  |  |  |
|  | Accuracy | Precision | Recall | F1_score | AUC | log_loss |  | Accuracy | Precision | Recall | F1_score | AUC | log_loss |
| Min | 0.724 | 0.315 | 0.161 | 0.213 | 0.528 | 9.399 | Min | 0.766 | 0.288 | 1.59E-04 | 3.18E-04 | 0.500 | 7.990 |
| Max | 0.728 | 0.334 | 0.168 | 0.223 | 0.533 | 9.528 | Max | 0.769 | 0.667 | 1.44E-03 | 2.87E-03 | 0.501 | 8.081 |
| Mean | 0.726 | 0.324 | 0.164 | 0.218 | 0.530 | 9.462 | Mean | 0.767 | 0.472 | 6.01E-04 | 1.20E-03 | 0.500 | 8.033 |
| Median | 0.726 | 0.323 | 0.164 | 0.217 | 0.530 | 9.460 | Median | 0.767 | 0.500 | 4.67E-04 | 9.33E-04 | 0.500 | 8.036 |
| STD | 1.10E-03 | 4.94E-03 | 2.06E-03 | 2.86E-03 | 1.36E-03 | 3.81E-02 | STD | 9.23E-04 | 1.10E-01 | 4.56E-04 | 9.09E-04 | 1.70E-04 | 3.19E-02 |

## Footnotes

1 https://github.com/scikit-learn

2 https://github.com/dmlc/xgboost

3 https://github.com/microsoft/LightGBM

4 https://github.com/catboost/catboost

5 https://github.com/tensorflow/tensorflow

6 https://github.com/scipy/scipy/blob/main/scipy/optimize/differentialevolution.py

7 https://github.com/scipy/scipy/tree/main/scipy/optimize

## Notes

### Competing Interest Statement

The authors have declared no competing interest.

### Funding Statement

This study did not receive any external funding.

### Author Declarations

This study has received ethical approval from The Australian Capital Territory Health Human Research Ethics Committee (Approval No. 2024/ETH01037).

